# The effect of eviction moratoria on the transmission of SARS-CoV-2

**DOI:** 10.1101/2020.10.27.20220897

**Authors:** Anjalika Nande, Justin Sheen, Emma L Walters, Brennan Klein, Matteo Chinazzi, Andrei Gheorghe, Ben Adlam, Julianna Shinnick, Maria Florencia Tejeda, Samuel V Scarpino, Alessandro Vespignani, Andrew J Greenlee, Daniel Schneider, Michael Z Levy, Alison L Hill

## Abstract

Massive unemployment during the COVID-19 pandemic could result in an eviction crisis in US cities. Here we model the effect of evictions on SARS-CoV-2 epidemics, simulating viral transmission within and among households in a theoretical metropolitan area. We recreate a range of urban epidemic trajectories and project the course of the epidemic under two counterfactual scenarios, one in which a strict moratorium on evictions is in place and enforced, and another in which evictions are allowed to resume at baseline or increased rates. We find, across scenarios, that evictions lead to significant increases in infections. Applying our model to Philadelphia using locally-specific parameters shows that the increase is especially profound in models that consider realistically heterogenous cities in which both evictions and contacts occur more frequently in poorer neighborhoods. Our results provide a basis to assess municipal eviction moratoria and show that policies to stem evictions are a warranted and important component of COVID-19 control.

## Introduction

The COVID-19 epidemic has caused an unprecedented public health and economic crisis in the United States. The eviction crisis in the country predated the pandemic, but the record levels of unemployment have newly put millions of Americans at risk of losing their homes [1–7]. Many cities and states enacted temporary legislation banning evictions during the initial months of the pandemic [8,9], some of which have since expired. On September 4th, 2020, the Centers for Disease Control and Prevention, enacting Section 361 of the Public Health Service Act [10], imposed a national moratorium on evictions until December 31st, 2020 [11]. This order, like most state and municipal ordinances, argues that eviction moratoria are critical to prevent the spread of SARS-CoV-2. It is currently being challenged in federal court (Brown vs Azar [12]), as well as at the state and local level [13,14].

Evictions have many detrimental effects on households that could accelerate the spread of SARS-CoV-2. There are few studies on the housing status of families following eviction [15,16], but the limited data suggest that most evicted households “double-up”--moving in with friends or family--immediately after being evicted (see Methods). Doubling up shifts the distribution of household sizes in a city upward. The role of household transmission of SARS-CoV-2 is not fully understood, but a growing number of empirical studies [17–22], as well as previous modeling work [23,24] suggest households are a major source of SARS-CoV-2 transmission. Contact tracing investigations find at least 20-50% of infections can be traced back to a household contact [25–28]. Household transmission can also limit or delay the effects of measures like lockdowns that aim to decrease the contact rate in the general population [18,23,29].

Here we use an epidemiological model to quantify the effect of evictions, and their expected shifts in household size, on the transmission of SARS-CoV-2, and the prospects of its control, in cities (Figure 1). We modify an SEIR (susceptible, exposed, infectious & recovered) model, previously described in Nande 2020 [23], to track the transmission of SARS-CoV-2 through a metropolitan area with a population of 1 million individuals. We use a network to represent contacts of the type that can potentially lead to transmission of SARS-CoV-2, between individuals grouped in households. We modulate the number of contacts outside the household over the course of the simulations to capture the varied effects of lockdown measures and their subsequent relaxation. We model evictions that result in ‘doubling up’ by merging each evicted household with one randomly-selected household in the network. In supplemental analyses, we examine what might happen if some proportion of evicted households enter homeless shelters or encampments.

**Figure 1:**
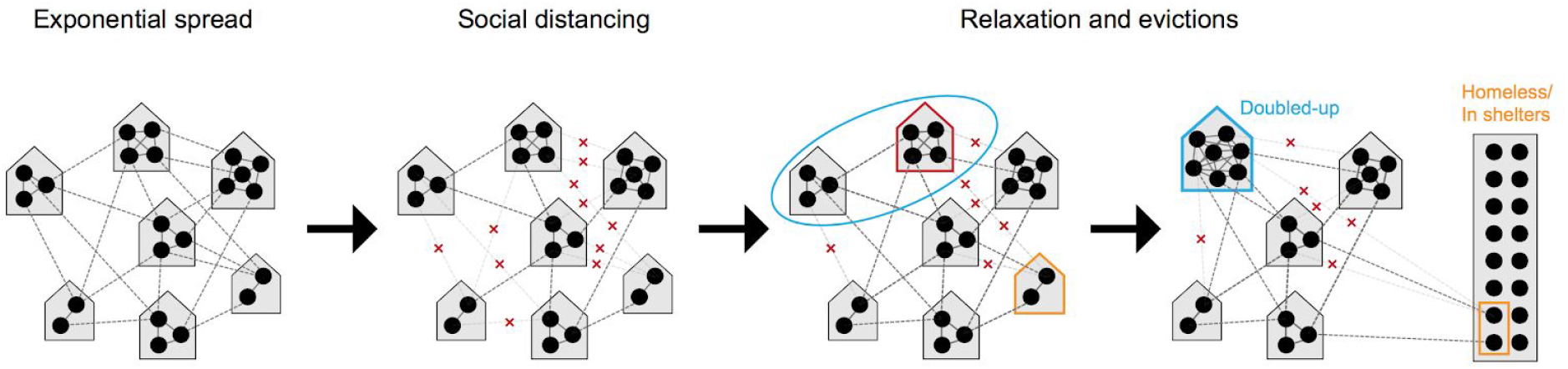
Modeling the effect of evictions on SARS-CoV-2 transmission. We model the spread of infection over a transmission network where contacts are divided into those occurring within a household (solid grey lines) versus outside the house (“external contacts”, dotted grey lines). Social distancing interventions (such as venue and school closures, work-from-home policies, mask wearing, lockdowns, etc) are modeled as reductions in external contacts (red x’s), while relaxations of these interventions result in increases in external contacts towards their baseline levels. When a household experiences eviction (red outline), we assume the residents of that house “double-up” by merging with another house (blue circle), thus increasing their household contacts. Evictions can also directly lead to homelessness (orange outline), and residence in shelters or encampments with high numbers of contacts

The model is parameterized using values from the COVID-19 literature and other demographic data (see Methods). The timing of progression between stages in our epidemiological model is taken from many empirical studies and agrees with other modeling work: We assume an ∼4 day latent period, ∼7.5 day serial interval and R_0_ ∼ 3 in the absence of interventions, and ∼1% infection fatality risk. Household sizes are taken from the US Census [30]; households are assumed to be well-mixed, meaning all members of a household are in contact with each other. Individuals are randomly assigned contacts with individuals in other households. The number and strength of these “external” contacts is chosen so that the household secondary attack rate is ∼0.3 [20], and the probability of transmission per contact is approximately 2.3-fold higher for households, compared to external contacts [19]. Our model naturally admits a degree of overdispersion in individual-level R_0_ values. Baseline eviction rates, which we measure as percent of households evicted each month, vary dramatically between cities at baseline, as do the expected increases due to COVID-19 (Supp. Figure 2, [31,32]). To capture this range of eviction burdens, we simulate city-wide eviction rates ranging from 0.1 - 2.0%.

Our initial models assume that mixing between households occurs randomly throughout a city, and that evictions and the ability to adopt social distancing measures is uniformly distributed. However, data consistently show that both COVID-19 and evictions disproportionately affected the same poorer, minority communities [33–39]. We therefore extend our model to evaluate the effect of evictions in a realistically heterogeneous city. We provide generic results, and then parameterize our model to a specific example--the city of Philadelphia, Pennsylvania.

Among large US cities, Philadelphia has one of the highest eviction rates. In 2016 (the last year complete data is available), 3.5% of renters were evicted, and 53% were cost-burdened, meaning they paid more than 30% of their income in rent [31,40]. In July, 2020 the Philadelphia city council passed the Emergency Housing Protection Act [41], in an effort to prevent evictions during the COVID-19 pandemic. The city was promptly sued by HAPCO, an association of residential investment and rental property owners [42]. Among other claims, the plaintiff questioned whether the legislation was of broad societal interest, rather than protecting only a narrow class (of at-risk renters). An early motivation behind this work was to assess this claim.

## Results

### Evictions drive increases in COVID-19 cases across cities

We simulate COVID-19 epidemic trajectories in single metro areas over the course of 2020. To recreate realistic scenarios in the model, we first collected data on COVID-19 cases and deaths aggregated for each US metropolitan statistical area with at least 1 million residents (∼50 cities). We then used hierarchical clustering to group the time series based on similar trajectories through Sept 2020, which resulted in four distinct groups of cities (Figure S3, Methods). Groups differed in the size of the spring wave, the degree of control in the early summer, and the occurrence and extent of a late summer wave (Figure S4-S8). For example, cities like New York, Boston, Philadelphia, and New Orleans (“Trajectory 1”) had large early epidemic peaks followed by dramatic and sustained reductions in cases; cities including Chicago, Baltimore, Seattle, and San Diego had substantial but smaller spring peaks that were controlled and then followed by a long plateau of cases over the summer (“Trajectory 2”); metros like St Louis, Raleigh, Salt Lake City, and San Francisco had much smaller spring outbreaks that were only partially controlled and then led to increases in the summer (“Trajectory 3”); and metros similar to Miami, Houston, Atlanta, and Phoenix experienced large mid-summer outbreaks (“Trajectory 4”). We calibrated our model to each of these four trajectory types by modulating the degree of reduction in external contacts over time (Methods, Table S1, and Figure S9).

Many local, county, and state-level eviction moratoria that were created early in the US epidemic were scheduled to expire in late summer 2020, so we modeled the effect of evictions taking place starting Sept 1 and continuing for the duration of the simulation. We assume that evictions happen at a constant rate per month, but that the backlog of eviction cases created during the moratoria results in 4 months worth of evictions occurring in the first month. We first considered an epidemic following Trajectory 1 (e.g. large spring epidemic peak followed by a strong lockdown and summer plateau, Figure 2). During fall 2020, we simulated a comeback of infection with a doubling time of 2-3 weeks, as has been observed across all large metros. By the end of 2020, 16% of individuals in our simulated scenario had caught COVID-19 in the absence of evictions. With a low eviction rate of 0.25%/month about 0.5% more of the population become infected compared to if there were no evictions. This increase corresponds to ∼5,000 excess cases per million residents. With a 1%/month eviction rate the infection level was ∼4% higher than baseline (Table 1). The exact values vary across simulations due to all the stochastic factors in viral spread considered in the model. These results highlight how the increased household spread that results from eviction-driven doubling-up acts synergistically with spread between members of different households during a growing epidemic to amplify infection levels.

**Figure 2:**
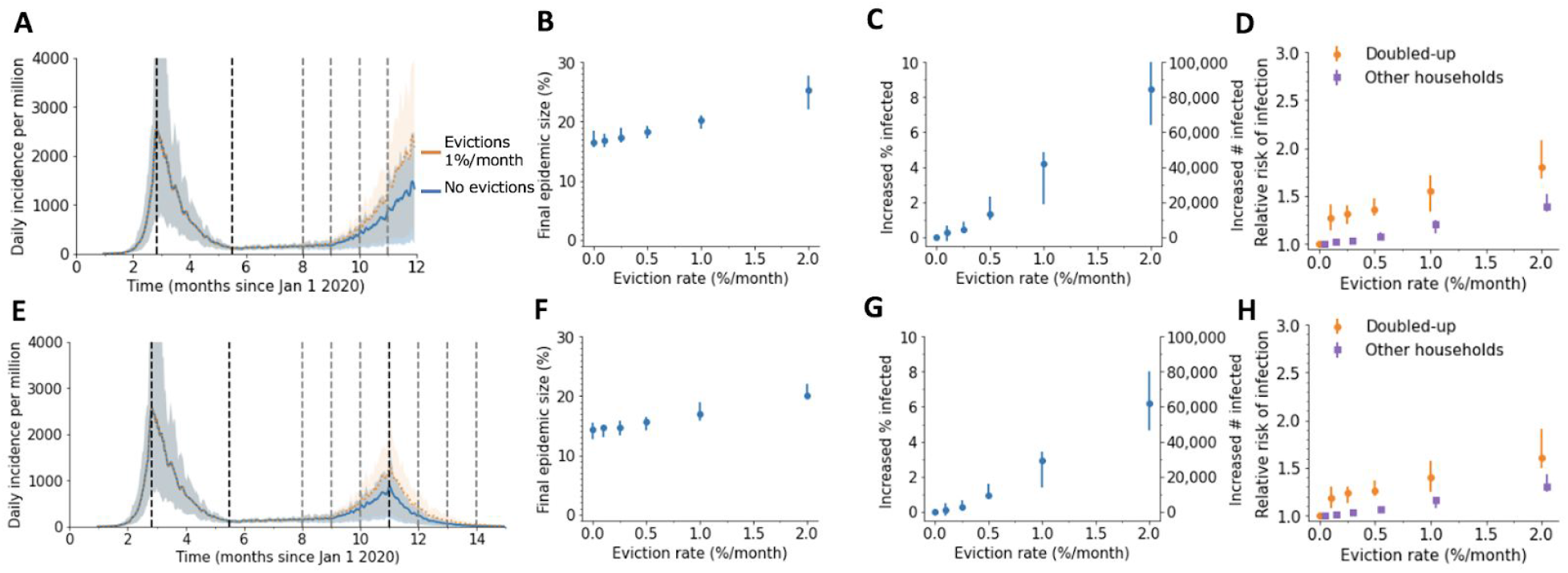
Impact of evictions on a SARS-CoV-2 comeback during Fall 2020. We model evictions occurring in the context of an epidemic similar to cities following “Trajectory 1”, with a large first wave and strong control in the spring, followed by relaxation to a plateau over the summer and an eventual comeback in the fall. Monthly evictions start Sept 1, with a 4-month backlog processed in the first month. A) The projected daily incidence of new infections (7-day running average) with and without evictions. Shaded regions represent central 90% of all simulations.The first lockdown (dotted vertical line) reduced external contacts by 85%, under relaxation (second dotted line) they were still reduced by 70%, and during the fall comeback they were reduced by 60% (fourth dotted line). B) Final epidemic size by Dec 31 2020, measured as percent of individuals who had ever been in any stage of infection. C) The predicted increase in infections due to evictions through Dec 31 2020, measured as excess percent of population infected (left Y-axis) or number of excess infections (right Y-axis). Error bars show interquartile ranges across simulations. D) Relative risk of infection in the presence versus absence of evictions, for individuals who merged households due to evictions (“Doubled-up”) and for individuals who kept their pre-epidemic household (“Other households”). E)-F) Same as above but assuming a second lockdown is instituted on Dec 1, and maintained through March 2021.

**Table 1.** Predicted excess SARS-CoV-2 infections when evictions are continued. Excess infections are measured as the increase in the cumulative percent of the population infected by Dec 31 2020 (or March 31 2021 if second lockdown implemented) if evictions resume on Sept 1, 2020, compared to if evictions were halted. Values are reported as median [IQR]. All results are from simulations of a metro area of 1 million individuals where evictions all result in “doubling up”. Corresponding epidemic trajectories are shown in Fig 2, 3. The US metropolitan statistical areas that roughly correspond to each scenario are shown in Figs S3, S5-S8.

| Trajectory | Cumulative prevalence w/o evictions | Eviction rate (%/month) |  |  |  |  |
| --- | --- | --- | --- | --- | --- | --- |
|  |  | 0.1% | 0.25% | 0.5% | 1% | 2% |
| Shown in Figure 2 |  |  |  |  |  |  |
| Trajectory 1 & comeback | 16.5<br>[15.7-18.5] | 0.25<br>[-0.24-0.63] | 0.46<br>[0.36-0.87] | 1.3<br>[1.0-2.3] | 4.2<br>[1.9-4.9] | 8.5<br>[6.4-10.8] |
| Trajectory 1 & lockdown | 14.3<br>[12.6-15.6] | 0.12<br>[-0.25-0.48] | 0.29<br>[0.23-0.64] | 0.90<br>[0.78-1.6] | 2.9<br>[1.4-3.4] | 6.2<br>[4.6-8.0] |
| Shown in Figure 3 |  |  |  |  |  |  |
| Trajectory 2 & comeback | 11.7<br>[10.3-12.5] | 0.38<br>[0.09-0.80] | 1.2<br>[0.67-2.2] | 1.4<br>[0.92-2.6] | 4.3<br>[3.1-5.3] | 9.8<br>[7.2-10.3] |
| Trajectory 2 & lockdown | 9.1<br>[7.8-10.1] | 0.18<br>[0.05-0.52] | 0.80<br>[0.43-1.4] | 0.93<br>[0.61-1.7] | 2.9<br>[2.3-3.5] | 6.9<br>[5.0-7.3] |
| Trajectory 3 & comeback | 8.9<br>[7.2-10.0] | 0.41<br>[-0.39-0.80] | 0.76<br>[0.36-1.04] | 1.7<br>[1.0-2.0] | 4.1<br>[3.6-4.4] | 9.4<br>[8.7-10.8] |
| Trajectory 3 & lockdown | 6.6<br>[5.4-7.5] | 0.29<br>[-0.36-0.60] | 0.43<br>[0.30-0.70] | 1.2<br>[0.63-1.4] | 2.7<br>[2.5-2.9] | 6.6<br>[6.2-7.6] |
| Trajectory 4 & comeback | 9.4<br>[8.1-10.4] | -0.07<br>[-0.16-0.34] | 0.47<br>[0.23-0.83] | 1.1<br>[0.94-1.6] | 2.6<br>[2.4-3.1] | 7.2<br>[5.9-7.5] |
| Trajectory 4 & lockdown | 8.4<br>[6.9-9.2] | 0.00<br>[-0.09-0.24] | 0.33<br>[0.18-0.64] | 0.89<br>[0.76-1.2] | 2.0<br>[1.9-2.3] | 5.9<br>[4.6-6.1] |

Our model predicts that even for lower eviction rates that don’t dramatically change the population-level epidemic burden, the individual risk of infection was always substantially higher for those who experienced eviction, or who merged households with those who did, compared to individuals whose households did not change (relative risk of infection by the end of the year, ∼1.28 [1.21, 1.37]). However, the increased risk of infection was not only felt by those who doubled-up: even for individuals who were neither evicted nor merged households with those who did, the risk of infection compared to the counterfactual scenario of no evictions was 1.04 for an eviction rate of 0.25%/month and 1.4 for 2.0% evictions per month (Figure 2D). This increased risk highlights the spillover effects of evictions on the wider epidemic in a city.

We then considered the same scenario but assumed the epidemic resurgence was countered with new control measures imposed on Dec 1. The epidemic was eventually controlled in simulations with or without eviction, but the decline was slower and the intervention less effective at reducing epidemic size when evictions were allowed to continue (Figure 2E-H). Following the epidemic until March 31 2021 at which point it was nearly eliminated locally, the final size was 0.3% greater with 0.25%/month evictions and 3% larger with 1%/month evictions. Larger households, created through eviction and doubling up, allow more residual spread to occur under lockdowns. Allowing evictions to resume will thus compromise the efficacy of future SARS-CoV-2 control efforts.

Cities across the US experienced diverse epidemic trajectories through August 2020, which we found could be summarized by three additional trajectory patterns (Figure 3, Figure S6-S8). These trajectory patterns, when used to calibrate our model, provide information on the prevalence of susceptible, infected, and previously-recovered (immune) individuals at the time evictions resume, as well how these cases may be distributed across households. We found that for all trajectories, evictions lead to significant increases in COVID-19 cases, with anywhere from ∼1,000 to ∼10,000 excess cases per million residents attributable to evictions when eviction rates are lower (e.g. 0.25%/month) and the fall comeback is reversed by a strong lockdown in December 2020, to ∼ 50-100,000 excess cases for higher eviction rates (e.g 2%) and unmitigated epidemics (Table 1). In most of these scenarios, there was ∼1 excess infection in the city attributable to each eviction that took place.

**Figure 3:**
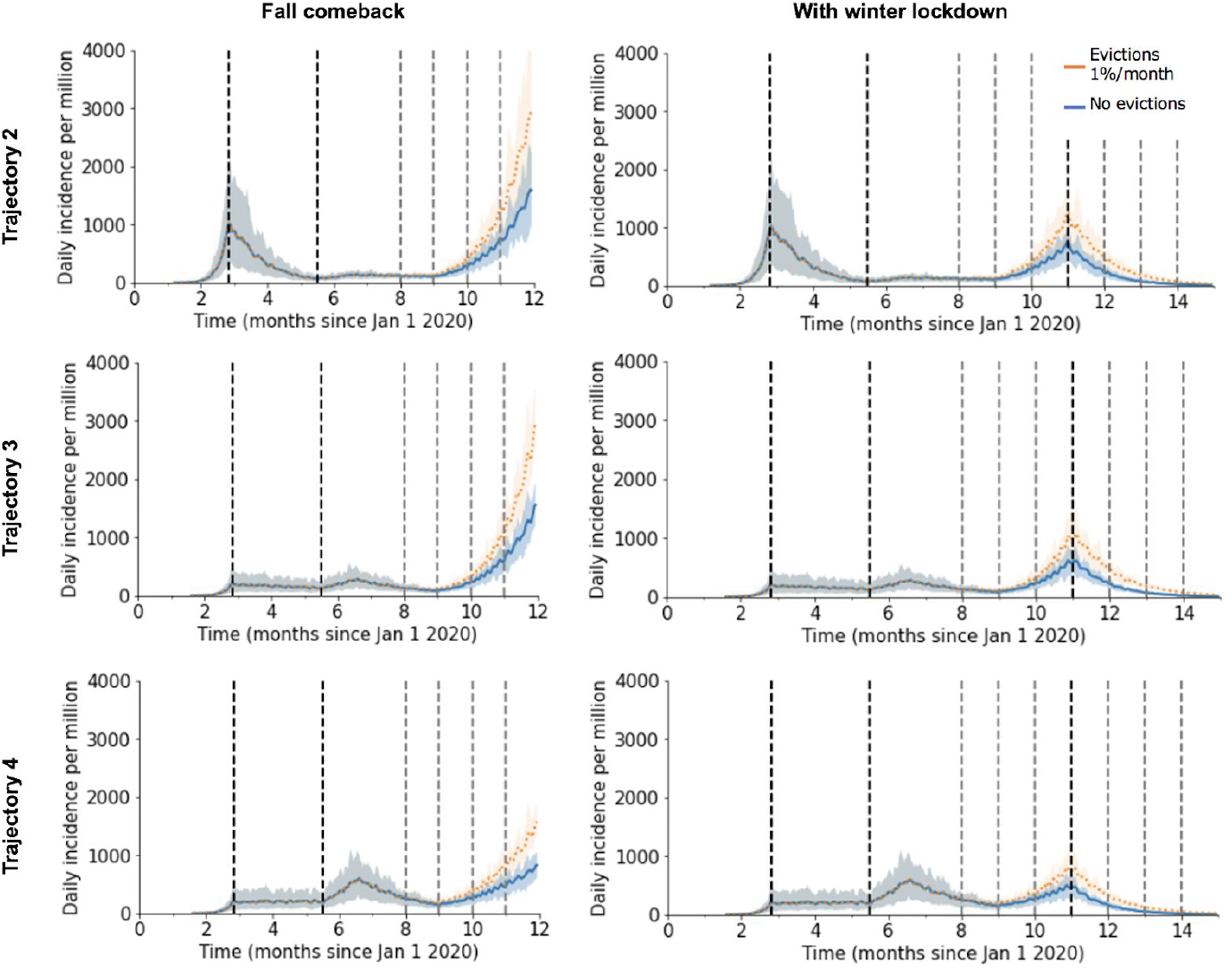
Alternate epidemic trajectories of SARS-CoV-2 before and after evictions in a large city. Each panel shows the projected daily incidence of new infections (7-day running average) with and without evictions at 1%/month with a 4 month backlog, starting on Sept 1 2020. Shaded regions represent central 90% of all simulations. In the left column, spread continues unabated through Dec 31 2020, whereas in the second column a new lockdown is introduced on Dec 1. Each trajectory scenario is created by calibrating the model to a group of US metropolitan statistical areas with similar patterns of spread (see Methods, Figure S5-S8). For all trajectory types, the degree of reduction in external contacts by control measures was modulated on dates March 25, June 15, July 15, and Oct 1, with values reported in Table S1.

We repeated the above simulations with different values of the household secondary attack rate (SAR) to check the sensitivity of our results to this value (Figure S10,11). As expected, when household transmission is more common (higher SAR of 0.5, Figure S10) evictions have a slightly larger effect on the epidemic, and when it is less common (lower SAR of 0.1, Figure S11), the effect of evictions is slightly smaller. If we assumed transmission risk from external contacts was equal to household contacts, but reduced the number of external contacts to maintain the same R_0_, results were similar to our baseline scenario (Figure S12). We also considered a scenario where evicted households always doubled-up with another household with whom they already had an external contact; results were similar (Figure S13). In a previous version of this work, completed before the ubiquity of the fall resurgence became apparent, we examined the impact of evictions under alternate fall scenarios, including a plateau of cases at September levels or more gradual increases [43]. We found that in general, evictions had less impact on infection counts when the epidemic was controlled and maintained at a constant plateau throughout the fall, however, a flat epidemic curve could still be associated with substantial detrimental effects of evictions if the incidence at the plateau is still relatively high.

### Evictions interact with urban disparities

Our results so far assume that every household in a city is equally likely to experience an eviction, and that SARS-CoV-2 infection burden and adoption of social distancing measures are homogeneously distributed throughout the population. In reality, evictions are concentrated in poorer neighborhoods with higher proportions of racial/ethnic minorities [39]. Similarly, individuals in these neighborhood types are likely to maintain higher contact rates during the epidemic due to high portions of essential workers among other reasons [44–47]. Many studies have shown higher burdens of infection and severe manifestations of COVID-19 in these demographic groups [33–38]. To examine the impact of city-level disparities on the interaction between evictions and disease spread (Figure 4), we simulated infection in cities consisting of two neighborhood types: a higher socioeconomic status (SES) neighborhood with no evictions and a high degree of adoption of social distancing measures (low contact rates), and a lower SES neighborhood with evictions, where the reduction in contact rates was less pronounced. We assumed that evicted households doubled-up with other households that are also in the low SES neighborhood (see Figure S15 for case when doubling-up can happen across neighborhoods). The epidemic time course was simulated using the same population-average transmission rates for each phase as for Trajectory 1 (Figure 2). In the absence of evictions, infection prevalence differed substantially by neighborhood (Figure 4B), and despite simulating the same overall reduction in contacts, the epidemic burden was higher when residual contacts were clustered in the poorer neighborhood (Figure 2B vs Figure 4D).

**Figure 4:**
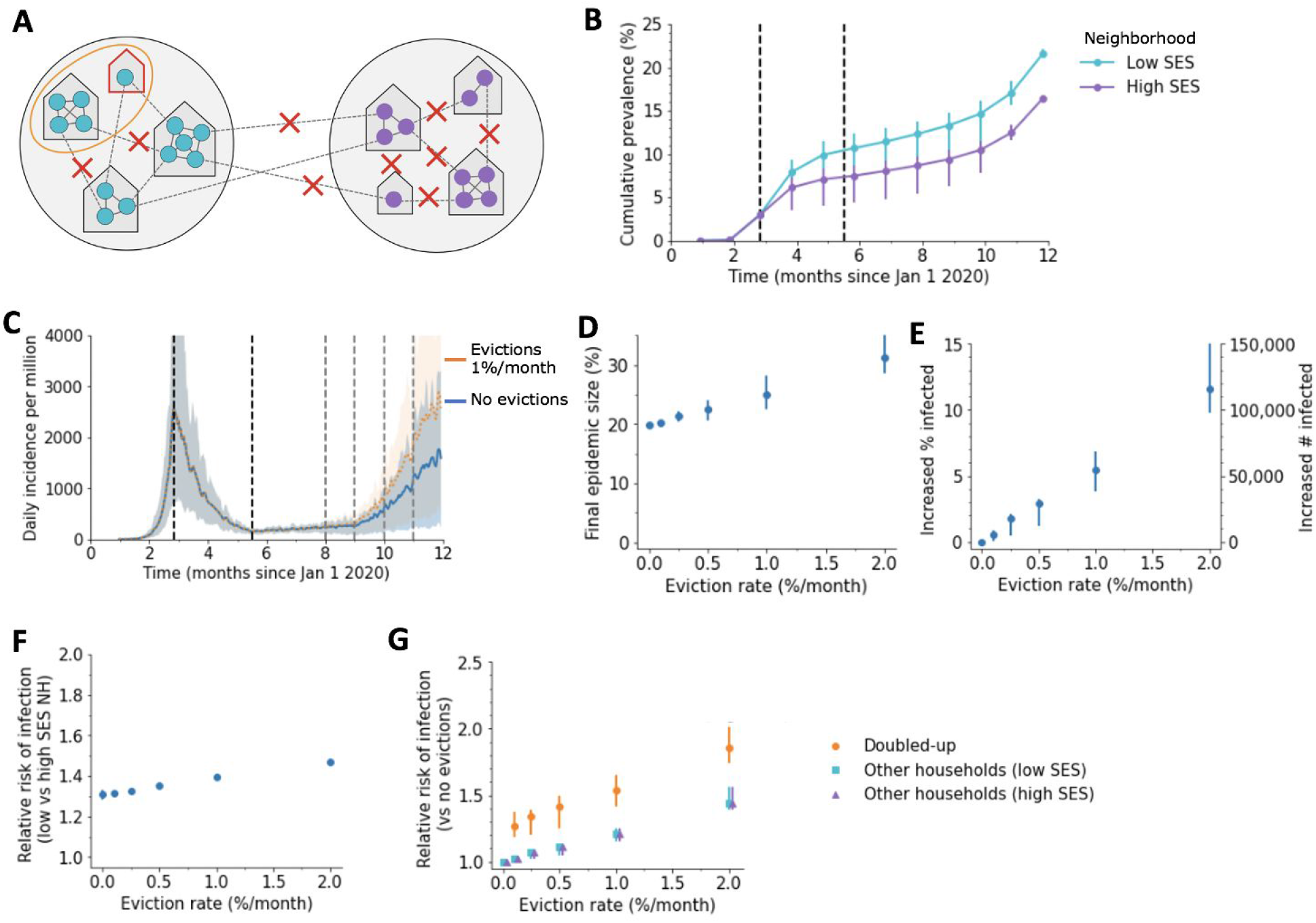
Impact of evictions on COVID-19 epidemics in heterogeneous cities. A) Schematic of our model for inequalities within a city. The city is divided into a “high socioeconomic status (SES)” (purple) and a “low SES” (teal) neighborhood. Evictions only occur in the low SES area, and individuals living in this area are assumed to be less able to adopt social distancing measures, and hence have higher contact rates under interventions (90% vs 80% reduction in external contacts during lockdown for 85% overall, 75% vs 65% during relaxation for 70% overall, and 65% vs 55% during fall comeback for 60% overall). Before interventions, residents are equally likely to contact someone outside the household who lives within vs outside their neighborhood. B) Cumulative percent of the population infected over time, by neighborhood, in the absence of evictions. C) The projected daily incidence of new infections (7-day running average) with 1%/month evictions vs no evictions. Shaded regions represent central 90% of all simulations. D) Final epidemic size by Dec 31 2020, measured as percent individuals who had ever been in any stage of infection, for the heterogenous city as compared to a homogenous city with same effective eviction rate and intervention efficacy. E) The predicted increase in infections due to evictions through Dec 31 2020, measured as excess percent of population infected (left Y-axis) or number of excess infections (right Y-axis). Error bars show interquartile ranges across simulations. F) Relative risk of infection by Dec 31 2020 for residents of the poor vs rich neighborhood. G) Relative risk of infection by Dec 31 2020 in the presence vs absence of evictions, for individuals who merged households due to evictions (“Doubled-up”) and for individuals who kept their pre-epidemic household (“Other households”).

In this heterogeneous context, we found, for equivalent overall eviction rates, larger impact of evictions on COVID-19 cases than if they had occurred in a homogeneous city. For example, by the end of 2020, for a low eviction rate (0.25%/month) we estimate 1.7% excess cases attributable to evictions in the heterogeneous model, as compared to 0.5% in the homogeneous version described above. For the higher eviction rate (1%/month) we estimate ∼5.4% excess infections due to eviction, as compared to ∼4.2% in the homogeneous model. Evictions also serve to exacerbate pre-existing disparities in infection prevalence between neighborhoods. For the hypothetical scenario we simulated, evictions increase the relative risk of infection for the low vs. high SES neighborhood from ∼1.3 to ∼1.5. However, due to spillover effects, individuals residing in the high SES neighborhood also experience an increased infection risk (up to 1.5-fold the no-evictions scenario) attributable to the evictions occurring in the other demographic group. These results hold even if we assume more extreme segregation between residents of each neighborhood (Figure S14) or if we allow evicted individuals to double up with residents of the high SES neighborhood (Figure S15), though the disparities across neighborhoods are more extreme in the former case and less extreme in the latter. Thus, our results in Table 1 may underestimate the impact of evictions on COVID-19 in realistically-heterogeneous US cities. The concentration of evictions in demographic groups with more residual inter-household transmission serves to amplify their effects on the epidemic across the whole city.

### Case study: Impact of evictions in Philadelphia, PA

Finally, we sought to combine these ideas into a data-driven case study motivated by the court case in Philadelphia, PA, mentioned above. In Philadelphia, like all major US cities, there is significant heterogeneity in housing stability and other socioeconomic factors that are relevant both to the risk of eviction and to COVID-19 infection [48,49]. An early study found clusters of high incidence of infection were mostly co-located with poverty and a history of racial segregation, such as in West and North Philadelphia [35]. A study of SARS-CoV-2 prevalence and seropositivity in pregnant women presenting to the University of Pennsylvania Hospital System found a more than 4-fold increase in seroprevalence among Black/non-Hispanic and Hispanic/Latino women, compared to white/non-Hispanic women, between April and June 2020 [37].

To include these important disparities in our modeling, we first used principal component analysis on a suite of socioeconomic indicators to classify zip codes in the city [50]. We obtained three zip code typologies: a higher income cluster, a moderate income cluster, and a low income cluster. The lower income cluster has both very high eviction rates and higher rates of service industry employment and essential workers. Properties of the sub-populations are summarized in Table 3 and detailed in Tables S1-S2.

We then translated these findings into our model by dividing the simulated city into three sub-populations. The fraction of external contacts of individuals residing in each cluster that were with individuals in each other cluster was estimated using co-location events measured by anonymous mobile phone data, which only includes interactions outside of homes and workplaces (see Table S5, Methods, Supplemental Methods). Individuals preferentially mix with others residing in the same cluster. The degree of adoption of social distancing measures, which differed for each sub-population and for each contact type, was assumed to be proportional to the reductions observed in the mobility data (Table S6). Cluster 1 experienced the largest reductions in mobility, followed by Cluster 3 then Cluster 2. Evicted households were merged with other households in the same sub-population. There are no datasets available tracking the geographic origin and destination of individuals experiencing evictions, but general housing relocations observed using the same mobility data were predominantly within the same cluster (Figure S16).

By tuning only the infection prevalence at the time that strong social distancing policies were implemented in March, we found that our model matched our best available information on the COVID-19 epidemic trajectory in Philadelphia (Figure 5). The simulated epidemic grew exponentially with a doubling time of 4-5 days until late March. Daily incidence of cases peaked shortly thereafter and deaths peaked with a delay of ∼ 1 month. Post-peak, new cases and death declined with a half-life of ∼ 3 weeks. In early June, control measures were relaxed, leading to a plateau in cases and deaths that lasted until early October. The seroprevalence over time predicted by the model was in general agreement with results from large serological surveys in different populations in Philadelphia or Pennsylvania as a whole [36,37,52,53], and our model predicted large disparities in seroprevalence among the clusters.

**Figure 5:**
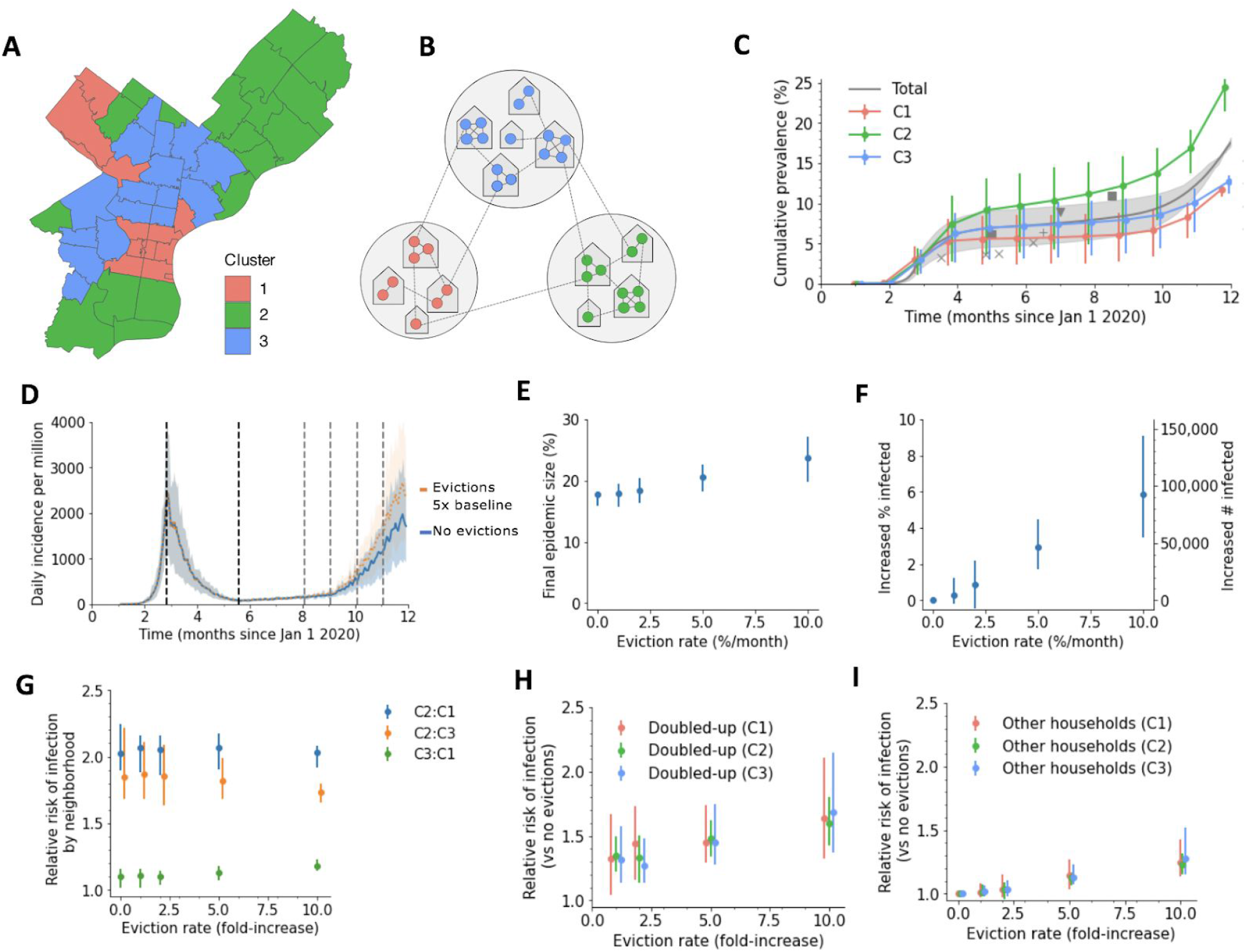
A detailed example of how evictions might affect SARS-CoV-2 transmission in the city of Philadelphia, Pennsylvania, USA. A) Map of Philadelphia, with each zip code colored by the cluster it was assigned to. Properties of clusters are Table 2 and Supplemental Tables 1-2 B) Schematic of our model for inequalities within the city. Each cluster is modeled as a group of households, and the eviction rate and ability to adopt social distancing measures vary by cluster. C) Simulated cumulative percent of the population infected over time, by cluster, in the absence of evictions. Data points from seroprevalence studies in Philadelphia or Pennsylvania: x [52], + [36], triangle [53], square [37]. D) The projected daily incidence of new infections (7-day running average) with evictions at 5-fold the 2019 rate vs no evictions. Shaded regions represent central 90% of all simulations. E) Final epidemic size by Dec 31 2020, measured as percent individuals who had ever been in any stage of infection. F) The predicted increase in infections due to evictions through Dec 31 2020, measured as excess percent of population infected (left Y-axis) or number of excess infections (right Y-axis). Error bars show interquartile ranges across simulations. G) Relative risk of infection by Dec 31 2020 for residents compared by neighborhood. H-I) Relative risk of infection by Dec 31 2020 in the presence vs absence of evictions, for individuals who merged households due to evictions (“Doubled-up”, H) and for individuals who kept their pre-epidemic household (“Other households”, I).

Simulations suggest that allowing evictions to resume could substantially increase the number of people with COVID-19 in Philadelphia by Dec 31, 2020, and that these increases would be felt among all sub-populations, including those with lower eviction rates (Figure 5). We predict that with evictions occuring at only their pre-COVID-19 rates, the epidemic would infect an extra 0.3% of the population (∼ 4700 individuals). However, many analyses suggest that eviction rates could be much higher in 2020 if allowed to resume, due to the economic crisis associated with COVID-19 (Figure S2, [32,51]). If eviction rates double, the excess infections due to evictions would increase to 0.9%; with a 5-fold increase in evictions, predicted by some economic analyses, this would increase to 2.6% or ∼53,000 extra infections. At this rate we estimate a 1.5-fold [1.3,1.6] increase in risk of infection for individuals in households that doubled-up, and a 1.1-fold [1.1,1.2] relative risk in other households, compared to the counterfactual situation in which a complete eviction moratorium was in place and enforced. Despite the differences in both baseline infection rates and eviction rates, all three sub-populations experienced similar proportional increases in infection levels due to eviction (Figure 5H,I). Overall our results suggest that eviction moratoria in Philadelphia have a substantial impact on COVID-19 cases throughout the city.

**Table 2:** Properties of neighborhood clusters in Philadelphia used in simulations.

|  | Population<br>in cluster<br>(%) | Eviction rate<br>at baseline<br>(%/month) | Estimated<br>essential<br>workers<br>(%) | Mobility<br>reduction<br>(%, June<br>2020) | Summary of cluster<br>characteristics |
| --- | --- | --- | --- | --- | --- |
| Cluster 1 | 17 | 0.07 | 0.30 | 89 | High income, high % renters, low % cost burdened, low % <age 18, low % female headed households, high mobility rate, high % white/Asian |
| Cluster 2 | 38 | 0.12 | 0.50 | 72 | Intermediate income, high % >65, low % renters, high cost burdened renters, high % foreign born, racial/ethnically diverse |
| Cluster 3 | 45 | 0.21 | 0.54 | 82 | High poverty, low income, high % renters, high vacancy, high % female headed households, high service workers, high % Black/Hispanic |
| Total | (1.58<br>million) | 0.15 | 0.48 | 79 |  |

## Discussion

Our analysis demonstrates that evictions can have a measurable impact on the spread of SARS-CoV-2 in cities, and that policies to stem them are a warranted and important component of epidemic control. The effect of evictions on an epidemic is not limited to those who were evicted and those who received evicted families into their homes. Other households experienced an increased risk of infection due to spillover from the transmission processes amplified by evictions in the city.

The most immediate implication of our findings is their relevance to the continued debate taking place in US legislatures and courts over the fate of local and national eviction moratoria. Our results suggest that the CDC-mandated national order prohibiting evictions from Sept 4 - Dec 31 2020 likely prevented thousands of excess COVID-19 infections for every million metropolitan residents. Moreover, simulations show that allowing evictions would increase the relative risk of infection for all households, not just a narrow class of those experiencing eviction. Thus, the legislation is of broad societal interest. A federal judge, citing testimony based on an earlier version of this model, recently issued an opinion in favor of the city of Philadelphia’s eviction moratorium [42]. More generally, our simulations show that in the case of COVID-19, preventing evictions is clearly in line with the mandate of the Public Health Service Act to invoke measures “necessary to prevent the introduction, transmission, or spread of communicable diseases” [10].

Across all scenarios evictions aggravated the COVID-19 epidemic in cities. The effect was greatest in scenarios when the epidemic was growing rapidly during the time evictions took place, but evictions also significantly increased the number of individuals infected under strong control measures. Even results of models that assume a homogenous population show significant effects of evictions on the city-wide prevalence of SARS-CoV-2 infection. However, when we modeled a heterogeneous city, the effect was more pronounced--presumably because the increase in household size is more concentrated, and those sections of the city affected by evictions, more connected.

The fates of households that experience eviction is difficult to track. Existing data suggest that historically the majority of evictions have led to doubling up (e.g. Fragile Families Study [54]) and we therefore chose to focus our modeling on this outcome. However, the current economic crisis is so widespread it is unlikely that other households will be able to absorb all evicted families if moratoria are revoked, and thousands of people could become homeless, entering the already-over-capacity shelter system or encampments [6,7,55–57]. Shelters could only increase the impact of evictions on cases of COVID-19. The risk of contracting COVID-19 in homeless shelters can be high due to close contact within close quarters, and numerous outbreaks in shelters have been documented [7,58]. A recent modeling study suggests that, in the absence of strict infection control measures in shelters, outbreaks among the homeless may recur even if incidence in the general population is low and that these outbreaks can then increase exposure among the general population [59]. When we assigned even a small proportion of evicted households to a catchall category of shelters and encampments (Figure S17), with an elevated number of contacts, evictions unpredictably gave rise to epidemics within the epidemic, which then, predictably, spread throughout the city. These results are qualitatively in line with the effects of other high-contact subpopulations, such as those created by students in dorms [60,61] or among prisoners [62–64].

Other modeling studies have investigated ‘fusing households’ as a strategy to keep SARS-CoV-2 transmission low following the relaxation of lockdowns [23,65,66]. The strategy has been taken up by families as a means to alleviate the challenge of childcare among other issues [67,68]. Indeed there are conditions, especially during a declining epidemic, that fusing pairs of households does not have much effect on infection on the population scale [23]. While these findings are not in discord with our work, a voluntarily fused household is a very different entity than an involuntarily doubled-up household. While a fused household can separate in subsequent periods of higher transmission, a doubled-up household would likely not have such an option.

All uninfected families are alike; each infected family is infected in its own way. Our model simplifies the complex relationships within households which might affect the risk of ongoing transmission within a home, as well as the complex relationships that might initially bring a pathogen into a home (e.g. [69]). Relatedly, we choose a constant household secondary attack rate. A number of analyses suggest that the average daily risk to a household contact might be lower in larger households [17,19,22]. We hesitate to generalize this finding to households in large US cities that face possible eviction. Empirical studies of secondary attack rate in households of different sizes, in the relevant population and with adequate testing of asymptomatic individuals, are badly needed. If they show that infection rate does not scale with household size, then our estimates on the expected effects of eviction would be too large. We also have not considered the effects of household crowding, nor the age structure of households--evicted and otherwise--which can influence outcomes in a network model such as ours [70,71]. We have not considered the effects of foreclosures and other financial impacts of the epidemic which will likely also lead to doubling up and potentially homelessness. There are likely more complicated interactions between COVID-19 and housing instability that we have not modeled, such as the possibility that COVID-19 infection could precipitate housing loss [57], that eviction itself is associated with worsening health [72,73], or that health disparities could make clinical outcomes of COVID-19 infection more severe among individuals facing eviction or experiencing homelessness [59,74,75]. Finally we note that our model is not meant to be a forecast of the future course of the epidemic, nor the political and individual measures that might be adopted to contain it. We limit ourselves to evaluating the effect of evictions across a set of scenarios, and we limit our projections to the coming months. We also do not consider the possible effects of re-infection, or other details that contribute to the uncertainty of SARS-CoV-2 epidemic trajectories.

We caution against using the precise numerical estimates from our model to infer the effects of eviction moratoria in particular locations. While we have attempted to use the best current estimates of epidemiological parameters for COVID-19 transmission and to calibrate our model to epidemic trajectories common across US cities, many uncertainties remain. For example, there is uncertainty in our estimates of the basic reproduction number, the serial interval, the duration of immunity, the infection fatality rate, the fraction of transmission occurring within vs outside of households, the distribution of external contacts across individuals and households and the manner in which they are reduced during different stages of the epidemic. These quantities may vary by setting and over time. The distribution of cases across demographic groups (e.g. age, race/ethnicity) is not considered in detail in our model, but may impact the output. Our analyses of different common COVID-19 epidemic trajectories and our sensitivity analyses using different parameter assumptions suggest our qualitative results are robust, but that the precise numbers depend on the details. Ideally, observational studies of households conducted in localities where evictions moratoria (temporarily) expired or were ignored could provide more direct evidence for the impact of evictions on COVID-19 transmission. No such studies exist to date. A recent preprint by Leifheit et al, taking a difference in difference approach at the state level, suggests there was some signal of increasing cases when moratoria ended [77]. However, there are likely other contemporaneous policy decisions affecting COVID-19 transmission that occured in states allowing local eviction moratoria to lapse, making the assignment of causality difficult.

Cities are the environment of an ever-increasing proportion of the world’s population [76], and evictions are a force which disrupts and disturbs them. Just as abrupt environmental change can alter the structure of populations and lead to contact patterns that increase the transmissibility of infectious agents [78], evictions change with whom we have the closest of contacts-- those inside the home-- and this change, even when it affects only a small proportion of a population, can significantly increase the transmission of SARS-CoV-2 across an interconnected city.

## Methods

### Modeling SARS-CoV-2 spread and clinical progression

We describe the progression of COVID-19 infection using an SEIRD model, which divides the population into the following stages of infection: susceptible (S), exposed (infected but not yet infectious) (E), infected (I), recovered and assumed to be immune (R), and deceased (D). This model was previously described in Nande et al [23] and is similar to many other published models [29,71,79–81]. All our code is available in a Github repository : https://github.com/alsnhll/COVID19EvictionSimulations. We assume that the average duration of the latent period is 4 days (we assume transmission is possible on average 1 day before symptom onset), the average duration of the infectious period is 7 days (1 day presymptomatic transmission + 6 days of symptomatic/asymptomatic transmission), the average time to death for a deceased individual was 20 days, and the fraction of all infected individuals who will die (infection fatality ratio, IFR) was 1%. The distribution of time spent in each state was gamma-distributed with mean and variance taken from the literature (see Supplemental Methods). Transmission of the virus occurs, probabilistically, from infectious individuals to susceptible individuals who they are connected to by an edge in the contact network at rate β.

### Creating the contact network

We create a two-layer weighted network describing the contacts in the population over which infection can spread. One layer describes contacts within the household. We divide the population into households following the distribution estimated from the 2019 American Community Survey of the US Census [30]. In this data all households of size 7 or greater are grouped into a single “7+” category, so we imputed sizes 7 through 10 by assuming that the ratio of houses of size n to size n+1 was constant for sizes 6 and above. We assume that all individuals in a household are in contact with one another. The second layer constitutes contacts outside the household (e.g. work, school, social). While the number of these external contacts is often estimated from surveys that ask individuals about the number of unique close face-to-face or physical contacts in a single day, how these recallable interactions of varying frequency and duration relate to the true effective number of contacts for any particular infection is not clear. Therefore, we took a different approach to estimating external contact number and strength.

We assumed a separate transmission rate across household contacts (β_HH_) and external contacts (β_EX_). To estimate the values of these transmission rates, and the effective number of external contacts per person, we matched three values from epidemiological studies of SARS-CoV-2. First, since the transmission probability of household contacts is more easily measured empirically than that for community contacts, we backed out a value of household transmission rate (β_HH_) that would give a desired value of the secondary attack rate (SAR) in households. We used a household SAR of 0.3, based on studies within the United States as well as a pooled meta-analysis of values from studies around the world [17,18,20]. Secondly, we assumed that the secondary attack rate for household contacts was 2.3-fold higher than for external contacts, based on findings from a contact tracing study [82]. We used this to infer a value for the transmission rate over external contacts (β_EX_). Finally, we chose the average number of external contacts so that the overall basic reproductive ratio, R_0_, was ∼3, based on a series of studies [83–85]. We did this assuming that the distribution of external contacts is negative binomially distributed with coefficient of variation of external contacts of 0.5, and using a formula for R_0_ that takes this heterogeneity into account [86]. We explore the effects of different assumptions surrounding R_0_, the household SAR, and the ratio between the transmission rate within and between households in sensitivity analyses, shown in the Supplement.

### Modulating contact rates to recreate epidemiological timelines

We assume that the baseline rate of transmission across external contacts represents the value early in the epidemic, before any form of intervention against spread (e.g. workplace or school closures, general social distancing, masking wearing). To recreate the trajectories of COVID-19 in U.S. cities throughout 2020, we instituted a series of control measures and subsequent relaxations in the simulations at typical dates they were implemented in reality. In the model, these modulations of transmission were encoded as reductions in the probability of transmission over external contacts. The timing and strength of these modulations in external contacts were chosen in a model calibration procedure that involved first clustering U.S. metro areas into groups based on common epidemic time courses and then tuning the model to recreate a trajectory typical of each group (described below, see Figure S3-S8, Table S1). For the simulations involving cities divided into multiple neighborhoods, we represented disparities by allowing the strength of these control measures to differ depending on the origin and destination neighborhood of each external contact. For the theoretical two-neighborhood city (Figure 4), individuals in the higher SES neighborhood were assumed to be better able to adopt social distancing measures and thus were modeled with larger reductions in external contacts. For Philadelphia, the reductions were informed by mobility data which tracked co-locations occurring between residents of different zipcodes (see Details below).

### Categorizing city-specific COVID-19 trajectories

To estimate the impact of evictions on COVID-19 transmission, it is necessary to realistically simulate the state of the epidemic at the time evictions are occuring. Theoretically, this would require estimates of infection prevalence over time, the distribution of infections across households, real-time estimates of contact rates of infectious individuals, as well as knowing how the epidemic would unfold in the future. Since in general we don’t have direct estimates of these quantities, our approach was to create realistic scenarios by simulating the entire epidemic course before evictions began, reproducing the trajectories seen in data. Then, for forward projections (which in our case is Sept 1 2020 onwards), we considered alternate scenarios of epidemic control and infection resurgence.

Our simulation is at the level of a single U.S. metropolitan area. To avoid the extreme computational costs of fitting a stochastic network model to every U.S. city of interest and including all the observational processes that affect real data (like testing rates, reporting delays, etc), we take a simplified model calibration approach. First, we collected county-specific daily COVID-19 case and death reports from the New York Times database (which is based on reports from state and local health agencies)[87], and aggregated these into metropolitan statistical areas (MSAs). Then, we chose all metropolitan areas with at least 1 million residents (53 cities) and used dynamical time warping and hierarchical clustering (using the `hclust` function with method `ward.D` and the `dtw` package in R) on case and death time courses (normalized by population) up to Aug 31 2020 to group cities with similar trajectories. Four groups naturally emerged from this analysis (Figure S3), representing at one extreme metros like New York City and Boston that had large first waves followed by a long period of control over the summer, and at the other extreme cities like Houston and Phoenix that had their epidemic peak over the summer. For all city groups, we observed that the epidemic could roughly be broken down into five phases with corresponding date ranges 1) early epidemic: pre-March 25, 2) lockdown: March 25-June 15, 3) relaxation: June 15 - July 15, 4) continued relaxation or increased control: July 15 - Oct 1, 5) fall wave: Oct 1 - end of 2020. We then created a trajectory with the model to match each city-group, where during each phase we chose a degree of reduction in external contacts using a simple binary search such that the growth rate of cases or deaths was within the range observed for cities in that group (Table S1). The time between initial epidemic seeding (with 10 individuals) and the first lockdown was chosen to reproduce the size of the first wave. Because of issues with testing limitations and reporting mild or asymptomatic infections, the fraction of cases reported has most likely changed dramatically over the course of 2020, and so we prioritize recreating trajectories of deaths (vs cases) along with an assumed 1% IFR to infer total cases (Figure S9).

### Modeling the impact of evictions

We initially assume all evictions result in “doubling up”, which refers to when an evicted household moves into another house along with its existing inhabitants. There are very few studies reporting individual-level longitudinal data on households experiencing evictions [15,88–90], and none of those (to our knowledge) have published reports of the fraction of households that experience each of the possible outcomes of eviction (e.g. new single-family residence, doubling-up, homeless, etc). The best evidence available on doubling-up comes from the Fragile Families and Child Wellbeing Study, one of the only large-scale, longitudinal studies that follows families after eviction [15,16]. The study sampled 4700 randomly selected births from a stratified random sample of US cities >200,000 population [54], and has followed mothers, fathers and children from 1998 to the present. Among other topics, interviewers asked respondents whether they had been evicted, doubled up, or made homeless in the previous year. In four waves of available interviews among 4700 respondents in 1999-2001, 2001-2003, 2003-2006, and 2007-2010, 402 mothers reported being evicted in the previous year. Of these, an average of ∼65% reported having doubled up.

Based on the eviction rate, a random sample of houses are chosen for eviction on the first day of each month, and these houses are then each “merged” with another randomly-chosen household that did not experience eviction. In a merged household, all members of the combined household are connected with edges of equal strength as they were to their original household members. A household can only be merged once.

Homelessness is another possible outcome of eviction. However, a number of studies of the homeless population show that doubling-up often precedes homelessness, with the vast majority of the homeless reporting being doubled-up prior to living in a shelter [91–93]. In the Fragile Families Study data described above, only ∼17.4% reported having lived in a shelter, car or abandoned building. We subsequently model the minority of evictions that result directly in homelessness. These evicted households instead enter a common pool with other homeless households, and, in addition to their existing household and external connections, are randomly connected to a subset of others in this pool, mimicking the high contact rates expected in shelters or encampments. We consider 10% of evicted households directly becoming homeless, and the other 90% doubling up. Individuals are assigned to ‘shelters’ of size drawn from a Poisson distribution with mean 20 and are connected to all other individuals in that shelter only. This number is estimated from considering both sheltered and unsheltered people, which each represent about half of individuals experiencing homelessness [55]. Data suggesting the average size of shelters is ∼25 people, based on assuming ∼half of the ∼570K homeless on any given night are distributed over 12K US shelters, which is similar to a Lewer et al estimate for the UK (mean 34, median 21) [59]. Unsheltered individuals are expected to have less close, indoor contacts, bringing down the average, and the portion of unsheltered homeless may grow as shelters reach capacity [55]. In the model, these connections get rewired each month, since the average duration of time spent in a single shelter is ∼ 1 month [94]. Contacts among homeless individuals are not reduced by social distancing policies. As a result, during the time evictions take place, the individual-level R_0_ for the homeless is ∼1.5-fold the pre-lockdown R_0_ for the general population, similar to Lewer et al [59].

Evictions begin in our simulations on September 1, 2020 and continue at the first of each month at a rate which we vary within ranges informed by historical rates of monthly evictions in metropolitan areas and projected increases (0%, .25%, .5%, 1%, or 2%) (see justification of this range in Supplemental Methods). Note that the denominator in our eviction rate is the total number of households, not the number of rental units. We assume a backlog of 4 months of households are evicted immediately when evictions resume, corresponding to the months during which most local or state eviction moratoria were in place (May through August 2020).

### Extension of the model to incorporate heterogeneity in eviction and contact rates

To capture heterogeneities in cities we divide the simulated city (population size 1 million) in two equal sized interconnected subpopulations--one consisting of high socioeconomic status (SES) households, and the other with low SES households. Connections between households (external contacts) could occur both within the same neighborhood and between the different neighborhoods. We considered two types of mixing for external connections - homogeneous mixing where 50% of external connections are within one’s neighborhood and heterogeneous mixing where 75% of external connections are within one’s neighborhood creating a more clustered population. Households in the low SES subpopulation experience all of the evictions in the city, and these evicted households are doubled-up with households from the same subpopulation. To capture the higher burden of infection with COVID-19 consistently observed in poorer sections of cities [33–39], during intervention we down-weighted external contacts among the low SES subpopulation less than in the high SES subpopulation (Table S1).

### Application to the city of Philadelphia, Pennsylvania, USA

To estimate the impact of evictions in the specific context of Philadelphia, PA, we first developed a method to divide the city up into a minimal but data-driven set of subpopulations, and then encode this into the model (as described above), taking into account the different rate of evictions and of adoption of social distancing measures across these subpopulations.

To create the subpopulations and capture key aspects of heterogeneity in the city, we first extracted 20 socio-economic and demographic indicators for each zip code of the city from the 2019 US Census, and ran an unsupervised principal component analysis [50] to cluster the zip codes based on similarities. The analysis resulted in three typologies: Cluster 1, a higher income rental neighborhood, Cluster 2, a moderate income and working-class owner neighborhood, and Cluster 3, a low-income rental neighborhood (Supplemental Table 1-2). Using zip code-level eviction rates for 2016 sourced from Eviction Lab [31], we estimated 0.7% of households in Cluster 1, 0.12% of those in Cluster 2, and 0.21% of those in Cluster 3 faced eviction each month at baseline. We consider scenarios where these rates are increased 2, 5 and 10-fold during the pandemic.

We used mobility data provided by Cuebiq to estimate the degree of contact between residents of the different clusters and how it varied throughout the epidemic. Cuebiq uses data from mobile phone users who have opted-in to share location information with certain applications, and we defined contacts using co-location events where two individuals were in the same 8-character geohash for 15-minutes or more. All data was anonymous and corrected for degree of population sampling (see Supplementary Methods for details). The percent of all within-Philadelphia contacts that occur with residents of a particular cluster is shown in Table S5. Between ∼50-75% of contacts were with individuals living in the same cluster, whereas ∼15-35% occurred in any other specific cluster.

We then calibrated our model to match the best available information regarding the epidemic trajectory for Philadelphia. The goal was to create a trajectory that captured the size of the epidemic peak (measured by daily death counts), the growth rate or decline of cases during each major phase of the epidemic (early phase, post-lockdown, post-relaxation, fall comeback), and the observed seroprevalence at different timepoints [36,37,52,53]. The reduction in mobility over time, measured as the percent reduction in contacts between residents of a pair of clusters averaged over 1 month, was used to determine the reduction in external contacts during each epidemic phase. To do this, in simulations we imposed a strong lockdown (overall 90% reduction in contacts) on March 23, 2020 when the cumulative prevalence of infected individuals was approximately 3% of the total population. This intervention was relaxed on June 15, allowing cases and deaths to plateau over the summer (overall 77% reduction in contacts). We further relaxed measures (overall 66% reduction in contacts) on Oct 1 2020 to create a fall comeback. Since no changes in mobility were observed during this time despite large increases in cases observed during this time period, we hypothesized that other biological or behavioral factors not represented in mobility data must be responsible for this comeback, and implemented it in the model as 15% increase in β_EX_. During each phase, each of the three clusters obtained via the typology analysis had different strengths of social distancing measures in their interactions with each other cluster (Table S6).

Data on the fate of evicted households is extremely rare, and we do not know how likely evicted individuals would be to double-up with a household within their cluster or in another cluster. To estimate this, we created a cohort of individuals from the Cuebiq data who’s inferred home location had changed between Feb 2020 and Oct 202. Figure S16 reports the frequency of moves to and from each cluster. This data suggests that the vast majority of relocations were to other houses within the same cluster. Assuming that movement patterns following eviction would be similar, in the model we assumed that evicted individuals always double-up with another household in the same cluster.

## Data Availability

Data: All COVID-19 case and death data used in this study were downloaded from the publically-available New York Times repository in Github. Data on the distribution of US household sizes and on socio-demographic indicators of zipcodes in Philadelphia was obtained from the 2019 United States Census. Mobility data used in this study is available from Cuebiq through their Data for Good program. Restrictions apply to the availability of this data, which was used under license for the current study, and so are not publically available. All data aggregated at the level of zip-code clusters that was used in the models is presented in the Supplemental Tables. More detailed data is available from the authors upon reasonable request and permission from Cuebiq, and any researchers interested in working with the data can apply for an independent license from Cuebiq. Aggregated mobility metrics at the national, state, and CBSA level are publically available at https://covid19.gleamproject.org/mobility.
Code: All our simulation code is available in a Github repository. The repository also contains code for downloading and processing COVID-19 case and death data from the New York Times repository.

https://github.com/nytimes/covid-19-data

https://www.census.gov/data/tables/2019/demo/families/cps-2019.html

https://www.cuebiq.com/about/data-for-good/

https://github.com/alsnhll/COVID19EvictionSimulations

## Data Availability

All COVID-19 case and death data used in this study were downloaded from the publically-available New York Times repository in Github: https://github.com/nytimes/covid-19-data

Data on the distribution of US household sizes and on socio-demographic indicators of zipcodes in Philadelphia was obtained from the 2019 United States Census: https://www.census.gov/data/tables/2019/demo/families/cps-2019.html

Mobility data used in this study is available from Cuebiq through their Data for Good program (https://www.cuebiq.com/about/data-for-good/). Restrictions apply to the availability of this data, which was used under license for the current study, and so are not publically available. All data aggregated at the level of zip-code clusters that was used in the models is presented in the Supplemental Tables. More detailed data is available from the authors upon reasonable request and permission from Cuebiq, and any researchers interested in working with the data can apply for an independent license from Cuebiq. Aggregated mobility metrics at the national, state, and CBSA level are publically available at https://covid19.gleamproject.org/mobility.

## Code Availability

All our simulation code is available in a Github repository : https://github.com/alsnhll/COVID19EvictionSimulations. The repository also contains code for downloading and processing COVID-19 case and death data from the New York Times repository.

## Acknowledgments

This work was supported by grants from the US National Institutes of Health DP5OD019851 (ALH), R01AI146129 (MZL). MC and AV acknowledge support from COVID Supplement CDC-HHS-6U01IP001137-01 and from Google Cloud and Google Cloud Research Credits program to fund this project.

## Supplement

### Supplemental Methods

#### Model details and parameters

We simulate the spread of SARS-CoV-2 using a modified version of the standard SEIR (Susceptible, Exposed, Infectious, Removed) model that allows for gamma-distributed durations of infection and infection spread over a network of contacts. We estimate the duration of the latent period (E stage. time before onset of infectiousness) as 4 ± 4 days, based on an incubation period duration of 5 ± 4 days [95,96] and assuming 1 day of pre-symptomatic transmission (similar to other models [97,98], and based on estimates of the portion of transmission that is pre-symptomatic [99] and the observation that viral load peaks at/before time of symptom onset [100–102]). Then, we assume a serial interval distribution of 7.5 ± 5 days (measured in the absence of rapid case isolation or other controls) [85,95,103,104] and back out an infectious period duration of 7 ± 4 (length of I_1_ state). This value is also consistent with estimates of the duration of high-level viral shedding [100] and of the symptomatic phase of mild (non-hospitalized) infection [104–107]. These are the main parameters governing the relationship between the input value of R_0_ (which we use to back calculate the β’s) and the rate of early exponential growth rate observed in the simulation before any social distancing.

Although not the focus of this work, our model also offers the ability to track the progression to more serious clinical stages of infection as well as recovery (R) and death (D) [23]. Here we use these other infection stages (i.e. hospitalization, ICU stay) only to include realistic approximations for the distribution of the timing from symptom onset to death (∼20 ± 10 days, in agreement with [106,108,109]) and for the portion of infected individuals who may eventually die (∼1%, in agreement with [110–113]). Tracking deaths help us to recreate the trajectories of the epidemic across different US metropolitan areas (Figures 3, S3-8): Since seroprevalence surveys across the US have suggested that cases are massively underreported, we calibrate the model to reproduce the epidemic size in terms of death counts and use the IFR to infer total cases.

In results that report “seroprevalence”, this was extracted from the model as the fraction of all currently living residents who were in the R stage, which is a rough surrogate. For results reporting “final epidemic size” or cumulative prevalence, we counted all individuals who had ever been in any stage of infection (i.e. E, I, R, or D).

#### Calculating the secondary attack rate

The secondary attack rate (SAR) is defined as the fraction of contacts of an infected individual who are infected directly by them over the course of their infectious period. For an individual with infectious period of length T, the SAR is:

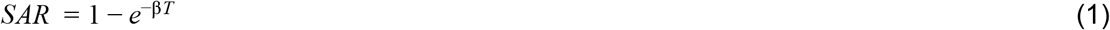

For a population of individuals in whom the infectious period is gamma distributed with mean *T* and shape parameter *k*, as in our model, the population-average SAR is :

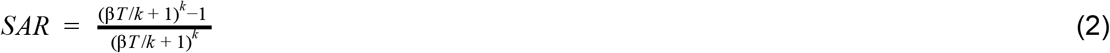

Considering transmission within households, we use observed values of the household SAR and our parameters of the infectious period (*T, k*) to back out the transmission rate within households:

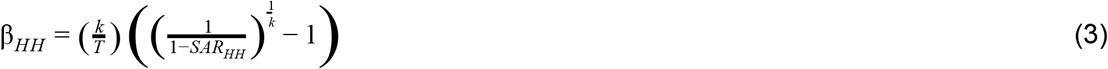

#### Estimating R_0_ for heterogeneous networks

We divide contacts down into two types - household and external - and similarly, the overall R_0_ can be decomposed into two components:

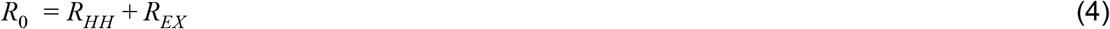

For a fixed uniform random network where everyone has *n* contacts, R_0_ can be approximated by:

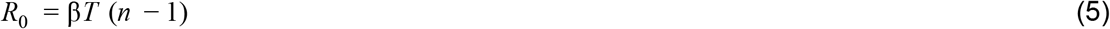

where the − 1 refers to the fact that an individual cannot infect the contact who infected them.

However, for random networks with significant variance in the number of contacts per individual, this formula substantially underestimates R_0_. Early in an epidemic, highly-connected individuals are more likely to be infected, and so R_0_ is more accurately estimated as:

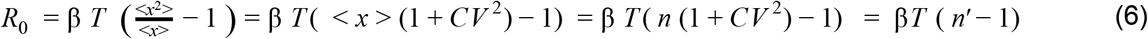

where *n* is the average degree, *CV* is the coefficient of variation, and *n*′ is an “effective” degree that takes into account the heterogeneity [86,114].

The variation in household sizes is small, so we use Eq 5 for R_0_^HH^, but we since we allow a large variation in external degree to account for realistic heterogeneity in human contact patterns and the degree of superspreading seen for SARS-CoV-2, we use Eq 6 for R_0_^EX^. The combined R_0_ is therefore:

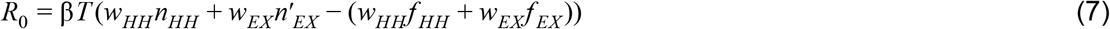

We define the weights of the household (external) layer *w*_*HH*_ (*w*_*EX*_) such that β_*HH*_ = β *w*_*HH*_ and β_*EX*_ = β *w*_*EX*_. The average contacts in each layer are *w*_*HH*_ and *w*_*EX*_, and the effective degree of the external layer (taking into account variance in connectivity) is 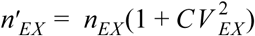. Instead of the − 1 term seen for R_0_ values for single layer networks (Eqs 5, 6), the − *w*_*i*_*f*_*i*_ value takes into account the fraction of infections caused by a particular contact type, with the terms given by 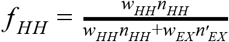 and *f* _*EX*_ = 1– *f* _*HH*_.

We define the weight of household contacts to be unity (*w*_*HH*_ = 1), so that the model value of β represents the rate of transmission over household contacts (β = β_*HH*_). Then we back out the weight of external contacts (*w*_*EX*_) from literature reports of the increased rate of transmissivity in households relative to outside (i.e. from reports of *w*_*EX*_ /*w*_*HH*_).

We can then use Eq 7 with fixed values of R_0_, T, w_HH_, w_EX_, n_HH_, and CV_EX_ to back out a value of n_EX_.

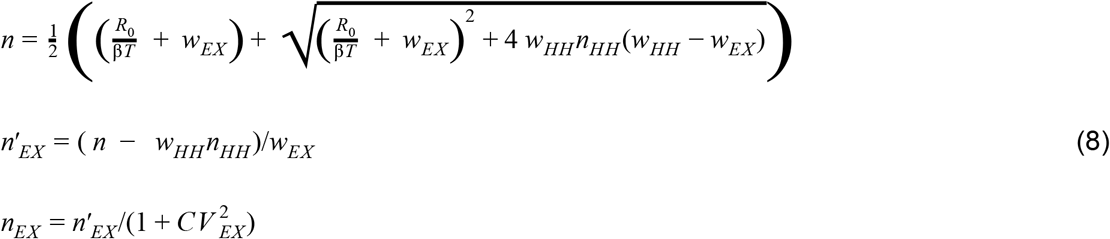

Based on the desired n_EX_ and CV_EX_, the parameters of a negative binomial distribution with parameters *p* (probability of success) and *r* (number of successes) are:

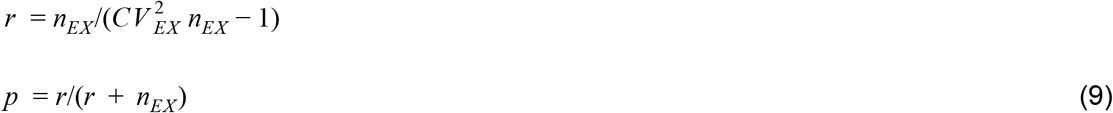

#### Estimating eviction rates across the US during the COVID-19 pandemic

To estimate the range of possible eviction rates across U.S. cities, we used data from Eviction Lab [31] and from an analysis by Stout [32]. Eviction rates are often expressed as rates per rental household so we scale the eviction rate per all households according to the percent of renter households in an area. Historically, across U.S. cities baseline evictions rates vary from ∼0.1%/month to ∼1%/month (Figure S2). However, high unemployment rates due to COVID-19 have already increased the potential eviction rate by creating a backlog of eviction filings throughout the country that could move forward quickly if current eviction moratoria were removed or struck down [7], and the continued high unemployment rates will only increase the eviction rate in metropolitan regions compared to their historical baselines.

To account for the uncertainty in the growth of eviction filings due to COVID-19 over the next year, we look at the fold-increase in unemployment in the same cities (compared to 2019) and assume that eviction rates (without any policies preventing evictions) could be increased by the same amount. Unemployment data was from the Bureau of Labor Statistics via the Department of Numbers (Figure S2A). In addition, we took estimates produced at a state-level by the consulting firm Stout, which used more detailed data on household income, savings, rent costs, unemployment, and recent national surveys (Figure S2B). These analyses suggest that eviction rates up to ∼2%/month are reasonable, though even higher rates are estimated with this method for some regions of the country. We consider the following eviction rates: 0% (comparison case), 0.1%, 0.25%, 0.5%, 1%, 2%/month which represent the spectrum of eviction rates for the majority of metropolitan regions.

#### Cluster specific down weighting of external contacts during intervention

##### Two clusters

In the case where the population was divided into two clusters - one for high socioeconomic status (SES) households and one for low SES households, there are three types of external connections that can occur. Connections between individuals belonging to high SES households (*x*_11_), those between low SES households (*x*_22_) and connections between one individual in a high SES household with one in a low SES household (*x*_12_). We reduce the weights of each type of contact (*r*_11_, *r*_22_, *r*_12_ < 1) such that the effective intervention efficacy of each cluster is reduced by the desired amounts (η_1_, η_2_< 1). The relationship between the reduction in weights of each type of contact and the effective intervention efficacy of each cluster can be calculated via the following system of equations,

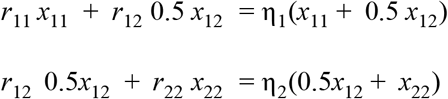

This system of equations is underdetermined as it has two equations and three unknowns (*r*_11_, *r*_22_, *r*_12_). We fix *r*_12_ = *r*_22_ = η_2_ and solve for 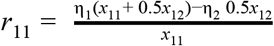.

#### Typological analysis of Philadelphia zip codes

Socio-economic indicators relevant to describing housing stability and COVID vulnerability were used to perform principal component analysis (PCA) and clustering to classify neighborhood types in the city of Philadelphia [50]. Data was collected by zip code tabulation areas from the 2019 US Census. The full list of indicators used and their value for each cluster is provided in Supplemental Table S3.

PCA and clustering resulted in three typologies (shown in Figure 5A): a higher income rental neighborhood, a moderate income and working class owner neighborhood, and a low income rental neighborhood. Cluster 1, the higher income neighborhood, is defined by high incomes and high housing costs. Cluster 2 is a slightly older, ownership centered neighborhood with lower poverty rates. Cluster 3, the lower income rental neighborhood, is defined by higher poverty rates, lower incomes, more children, and higher rates of service industry employment and essential workers. Details of these clusters are provided below. Considering the socio-economic stratification among these typologies, it’s likely that Type 3 zip codes will face higher levels of health and socio-economic vulnerabilities due to COVID-19. We then analyzed the population, household, and rental household tabulations, race, ethnicity, and eviction rates of each cluster, which were variables not included during the PCA.

To get the monthly eviction rate per household in Philadelphia for each cluster, we use eviction rate data reported for each zip code, from Eviction Lab [31]. Eviction Lab reports eviction rate per rental unit per year, and we used the reported fraction of all households that were renters (vs owners) to reframe the eviction rate as per all households per month.

##### Cluster 1

Cluster 1 is defined by higher housing costs and higher income when compared to the overall mean of the city. When compared to the overall mean, these zip codes have significantly higher median home values, higher gross rents, higher incomes, and a higher per capita income. They are predominantly rental units with lower rates of poverty and cost burdened households than the overall mean. Additionally, the percentage of residents who are essential workers is lower than the overall mean of the city. Considering the high housing costs, lower cost burdened rates, and lower rates of essential workers, it’s likely that these zip codes are economically stable and less vulnerable to the health and socio-economic impacts of COVID-19. When analyzing race and ethnicity of the clusters (see Supplemental Table S3), Cluster 1 zip codes are predominantly white with a small share of the population being residents of color. Cluster 1 zip codes also see the lowest eviction filing rates and eviction rates at 3.3% and 1.45% per rental household per year, respectively.

##### Cluster 2

Cluster 2 is defined by higher rates of homeownership and a slightly older population compared to the overall mean. Vacancy rates, poverty rates, and mobility rates are lower than the overall mean. When analyzing the overall means of all variables in this cluster (see Supplemental Table S1), the median household income is about equal to the overall mean and the median home value is just above the overall mean. Additionally, these zip codes have higher rates of essential workers among their residents when compared to the city’s overall mean. Considering the socio-economic conditions of this cluster, it’s likely that this type of neighborhood is moderate income and working class. Cluster 2 neighborhoods are far more diverse than cluster 1 neighborhoods (Supplemental Table S3), but still skew white with the overall mean of the white population in zip codes in cluster 1 being 51 percent. Cluster 2 sees a higher concentration of Black residents and a slight increase in the Latino population when compared to cluster 1. Zip codes in cluster 1 also see substantially higher eviction filing rates and eviction rates compared to cluster 1. The mean eviction filing rate and eviction rate are 7.8% and 3.8% per rental household per year, respectively.

##### Cluster 3

Cluster 3 represents lower income zip codes with higher rates of female headed households, higher poverty rates, higher vacancy rates, lower home values, and lower incomes when compared to the overall means of these indicators. These zip codes also have higher rates of residents employed in the service industry and as essential workers. This indicates that households in this type of neighborhood are more vulnerable to the health and socio-economic impacts of COVID-19. Zip codes in cluster 3 are predominantly nonwhite with higher Black and Latino populations than cluster 1 and 2. Eviction filing rates and evictions rates are also higher in cluster 3 zip codes than in other areas of the city with an eviction filing rate of 10.2% and an eviction rate of 4.7% per rental household per year.

#### Using mobility data to determine contact patterns and quantify social distancing in Philadelphia

Throughout the COVID-19 pandemic, aggregated mobile device data has been used for building and parameterizing epidemic models [29,115–118], as well as studying the impact of the pandemic on our mobility and social contacts [44,45,47,119,120]. Here, we use aggregated mobile phone data from more than 13,000 opted-in, anonymous users in Philadelphia, PA, USA to estimate the probability that residents of different zip codes (or zip code clusters) interact with one another in a given day based on co-location. This results in an estimated contact matrix where each entry corresponds to the average number of interactions that a resident of zip code *z*_*i*_ has with residents of zip code *z*_*j*_. We use data from Cuebiq, which provides data to academic and humanitarian initiatives through its Data for Good program (https://www.cuebiq.com/about/data-for-good/). These data are first-party and collected from anonymous users who have opted in to share their location data. The privacy of these users is further enhanced in several ways: First, users’ “personal areas” such as home locations are up-leveled to the Census block group level, which preserves key demographic properties while obscuring sensitive location information. Second, the measures derived from these data are again aggregated to the zip code level, further eliminating any identifiable information of the users. We have used these data with similar processing schemes in previous COVID-19-related papers [29,119,121,122].

#### Selecting a panel of users

In this study we consider mobility data from 13,333 users that have personal areas in the city of Philadelphia. These users were sampled from a larger panel of more than 5 million users nationwide. This original panel represents a subset of all active users in the Cuebiq dataset who met the following criteria: 1) users who were active in the dataset for at least 21 days during each month from January to June 2020, 2) users whose devices reported on average at least one location ping per hour, and 3) users with an average device geolocation accuracy of less than 50 meters.For more details about the criteria and composition of the panel, as well as the rationale for the selection criteria, see Klein et al. [119]. To see an interactive dashboard of different collective physical distancing measures that were computed with these data, see http://covid19.gleamproject.org/mobility.

#### Operationalizing opportunities for contact in mobile device data

We define an *opportunity for contact* as two devices being spatially co-located for some period of time. Here, spatial co-location is based on the longitude-latitude position of two devices, such that if two devices are within the same 8-character *geohash*, they qualify as being co-located. A geohash is a compressed string representation of the full longitude-latitude coordinates, and the more characters in a geohash, the finer scale resolution it will have; 8-character geohash are typically 25m^2^, with the largest dimensions being 19m x 38m at the equator [123]. Following guidance from the CDC about what constitutes a close contact, we treat co-location events of 15 minutes or more as a successful opportunity for contact. In sum: we define an opportunity for contact as a pair of devices being within the same 8-character geohash for 15-minutes or more.

#### Spatially-aggregated contact patterns across zip codes

In order to arrive at an aggregated cluster-to-cluster contact probability matrix, we first estimate a zip code-to-zip code contact matrix. This is done by first assigning each user to a “personal area” (i.e., an inferred home location—up-leveled to the zip code to preserve privacy—based on periods of nightly inactivity). Using these personal areas, we can estimate the likelihood that a resident of zip code *z*_*i*_ interacts with a resident of a nearby zip code *z*_*j*_ based on the sum of observed contact opportunities between users with personal areas of *z*_*i*_ and *z*_*j*_.

We re-weight observed contacts between users with personal areas in zip codes *z*_*i*_ and *z*_*j*_ in order to account for possible sampling bias in our panel of users. To do this, we first define *f*_*i*_ to be *n*_*i*_ */ N*_*i*_, which is the number of users with personal areas in the zip code, *n*_*i*_, divided by the total population of the zip code, *N*_*i*_. The adjusted estimate for the total number of contacts between a pair of zip codes *z*_*i*_ and *z*_*j*_ is then defined as *X*_*ij*_ *= c*_*ij*_ / (*f*_*i*_ *∗ f*_*j*_), where *c*_*ij*_ is the observed number of contacts between users with personal areas in zip codes *z*_*i*_ and *z*_*j*_. We normalize this to a per capita level within a given zip code by dividing by the population of that zip code, *x*_*ij*_ *= X*_*ij*_ */ N*_*i*_. When referring to “contacts” between a pair of regions (e.g. zip codes or clusters), we are referring to this quantity.

At the zip code level, we are now left with a 46 x 46 weighted contact matrix, ***X***_*τ*_, where *τ* denotes a time frame of interest. We define ***X***_*ref*_ to be the baseline contact matrix, corresponding to the average activity between January 16 and February 28, 2020, excluding holidays (as in Klein et al. [119]). Each week’s average contact patterns between pairs of zip codes can now be compared to the “typical” or baseline period, which was chosen because it represents activity after winter holidays and before large scale mobility disruptions were observed nationwide. Together, this lets us define the final matrix of interest, which describes the *percent of typical contacts between regions*. That is, elements *W*_*ij*_ of the matrix *W*_*τ*_ *=* ***X***_*τ*_/***X***_*ref*_ correspond to the percent of typical activity between zip codes *z*_*i*_ and *z*_*j*_. Lastly, we aggregate this into a cluster-to-cluster (3 x 3) matrix by taking the weighted sum of the elements of Wτ (zip code to zip code contacts); for every zip code in a given cluster, we add together the average contacts proportionally based on the typical total number of contacts, x_i_ = ***Σ***x_ij_.

Note that we exclude zip code 19112 from our analyses for privacy reasons due to its small population (estimates on the order of n=10 residents according to the 2019 ACS Survey).

#### Using mobile device data to estimate relocation patterns in Philadelphia

As mentioned above, we assume that users are more likely to relocate within the same cluster in the event of an eviction. We validate that assumption by studying the change in personal areas in our panel of users. In order to calculate this percentage, we recompute the (up-leveled) average nighttime location of the users in our panel during February and October, 2020. We use this position as a coarse estimate of the home zip code of these users. Among users whose home zip codes changed between February and October, we construct a matrix corresponding to the percent of relocations to each of the other zip codes in Philadelphia. In order to ensure user privacy, we only report percentages in this analysis.

The results of this analysis are shown in Figure S16, where each element of this “relocation matrix” represents the percent of users who started (in February) with a home zip code in a given cluster and ended up (in October) with a home zip code in another cluster. Note that the large majority of relocations take place within the same cluster. Users in Cluster 1 are more likely to relocate to other clusters than those in Cluster 2 and Cluster 3. While more systematic demographic analyses of these within-city relocation patterns are surely warranted, we use the results in Figure S16 only to parameterize our estimates of the rate of within- and between-cluster relocations for the epidemic simulations studied here.

## Supplemental Figures

**Figure S1:**
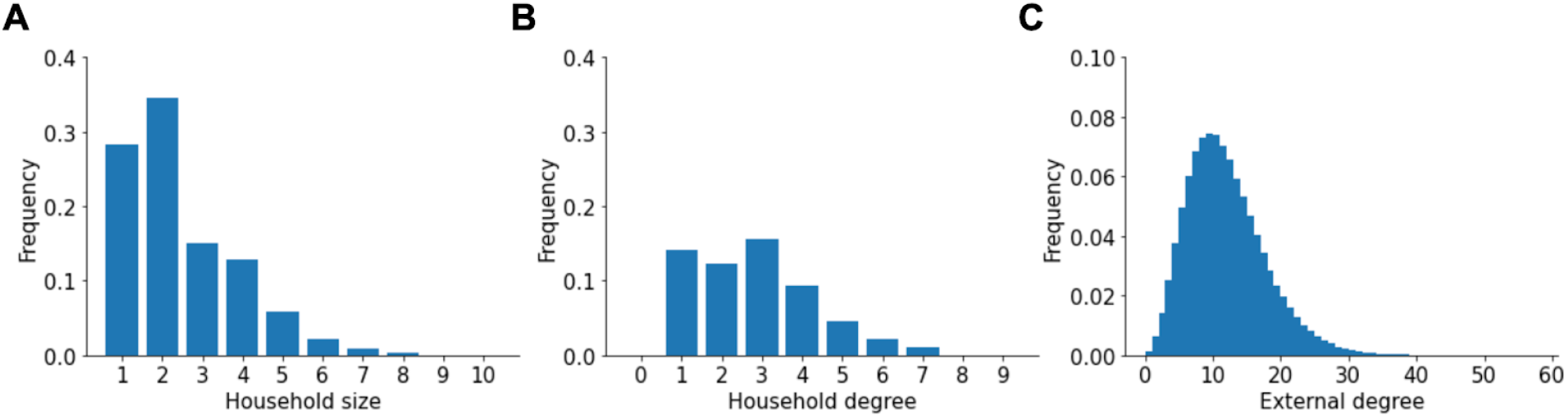
Degree distributions in the network. A) Distribution of household sizes. B) Distribution of the number of household contacts (degree). C) Distribution of the number of non-household (external) contacts.

**Figure S2:**
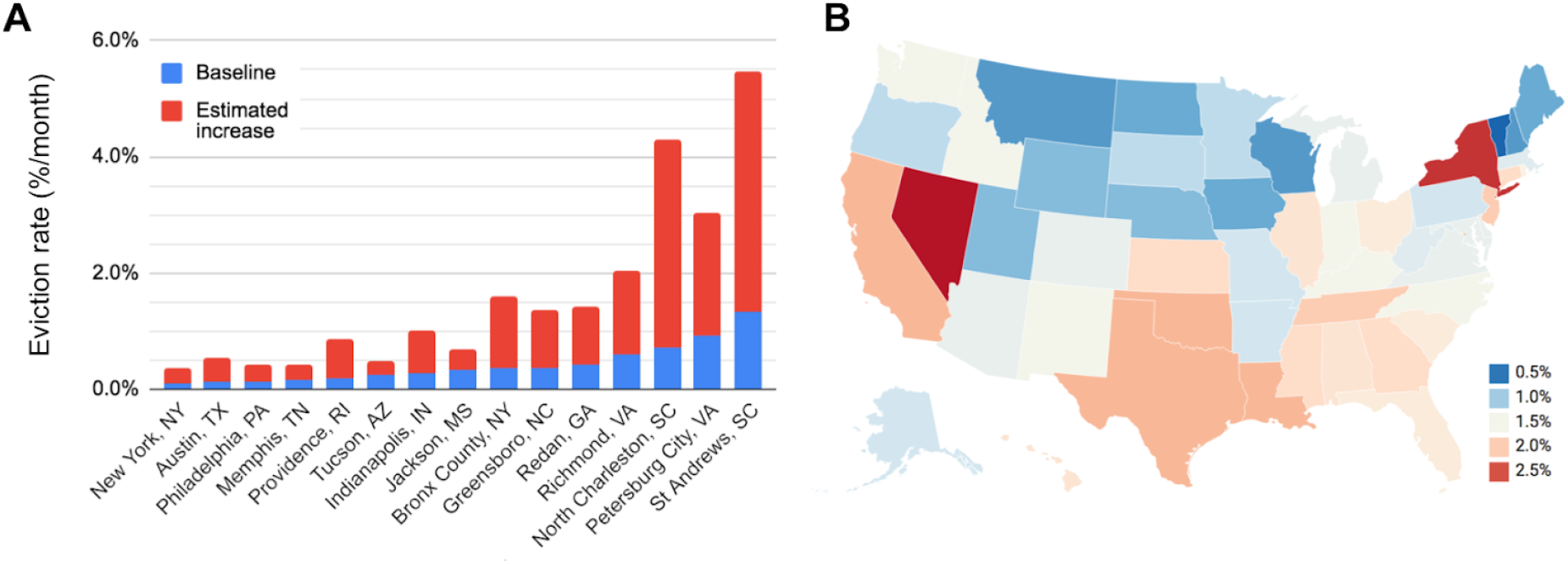
Baseline and estimated future eviction rates across selected US cities/districts and states. A) Baseline eviction rates (blue) and estimated increase in eviction rate based on increased unemployment rate (red). Data from Eviction Lab [31]. Rates are percent of all households experiencing eviction per month. In a city of ∼1 million people and a US-average household size of ∼2.5, an eviction rate of 0.1%/month corresponds to about 400 evictions per month, and a rate of 2%/month works out to 8000/month. These cities were chosen to represent diversity in size, geography, and eviction rates B) Estimates for the percent of households facing eviction per month over the next four month in each state. Values came from an analysis from consulting firm Stout [32] and were based on US census data and surveys.

**Figure S3:**
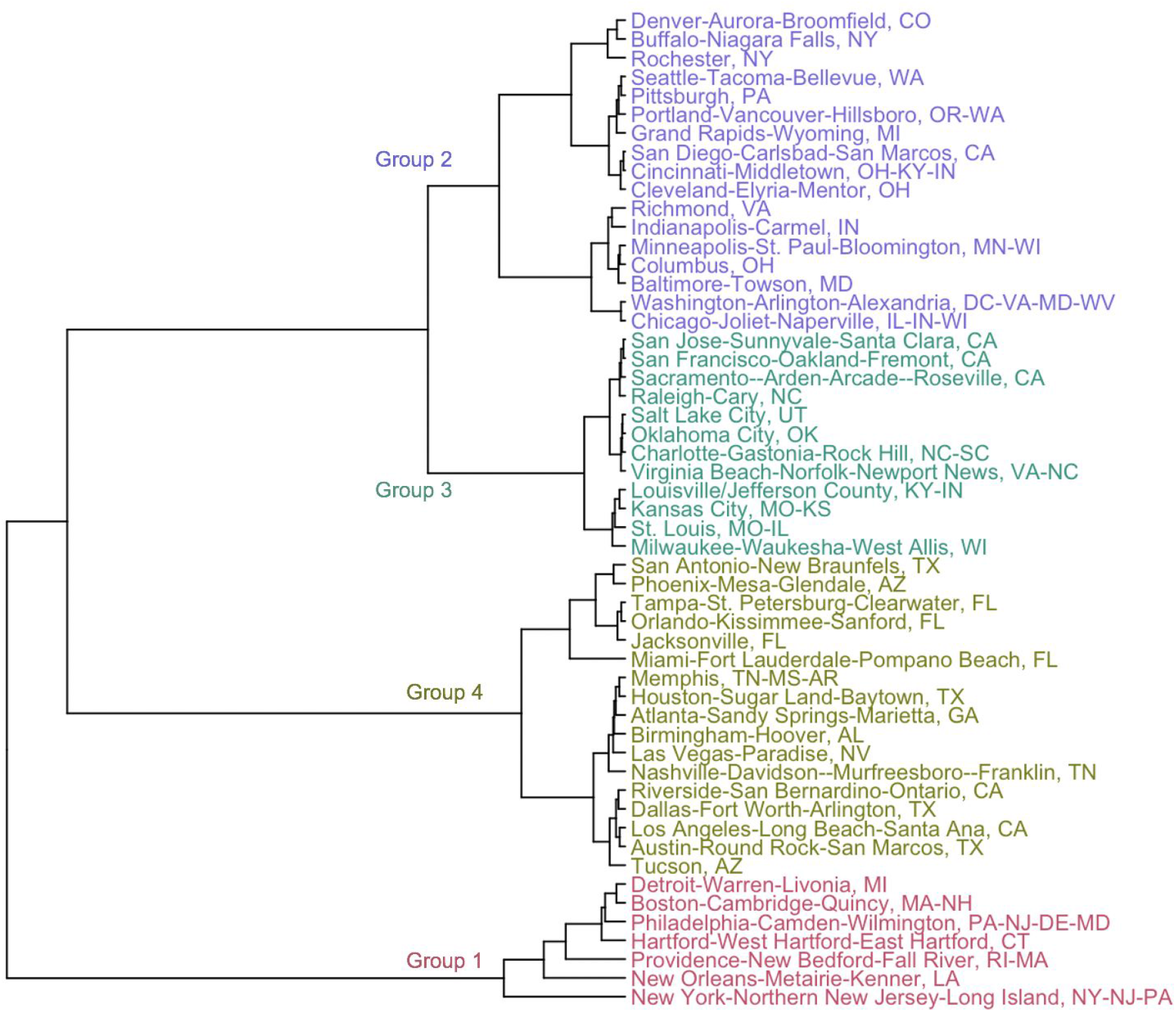
Clustering cities based on COVID-19 epidemic trajectories. Dendrogram from results of hierarchical clustering of US metropolitan statistical areas based on the trajectory of COVID-19 cases and deaths through Sept 1 2020. City names are colored based on group membership when four clusters were chosen. See Methods for details.

**Figure S4:**
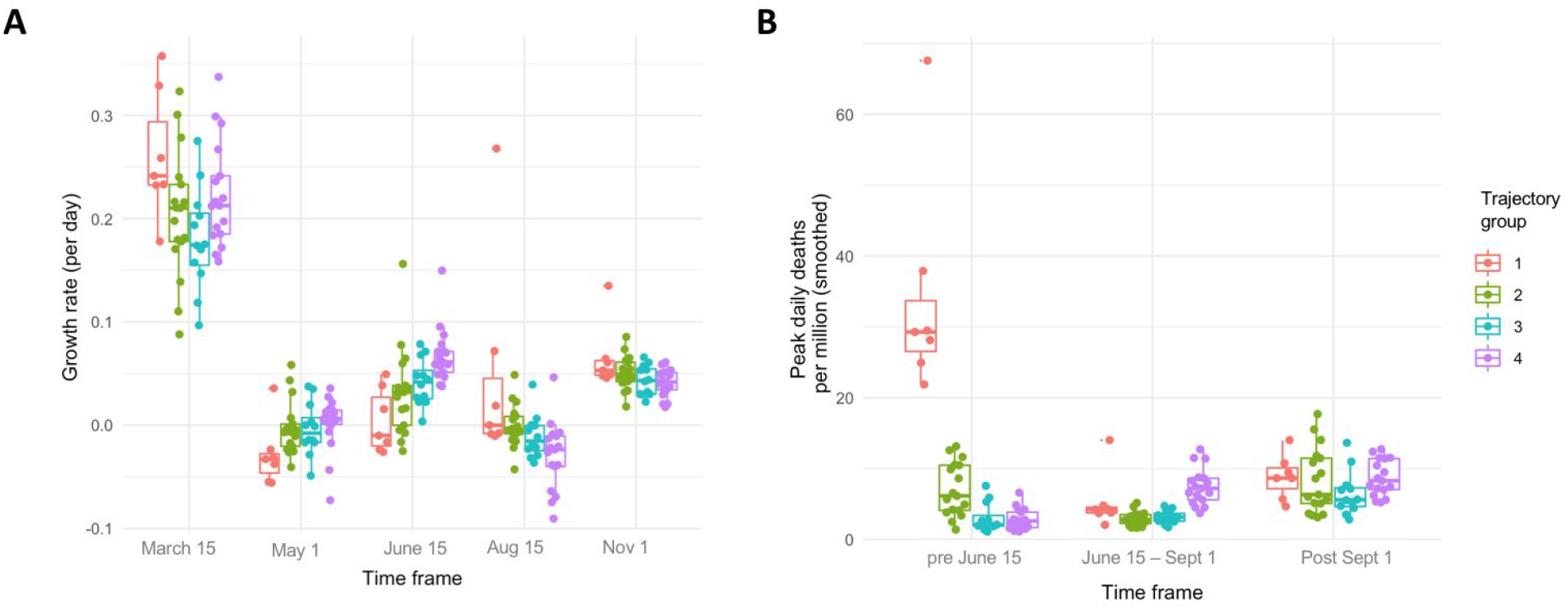
Characteristics of the epidemic trajectory in each group of cities. A) Growth rates cases, measured for two weeks starting from the stated date. B) Peak deaths observed in each stated date interval. Box plots show median, IQR, and 95th percentiles. All metrics were extracted from rolling 7-day averages of the daily incidence of new cases or deaths. See Methods for details.

**Figure S5:**
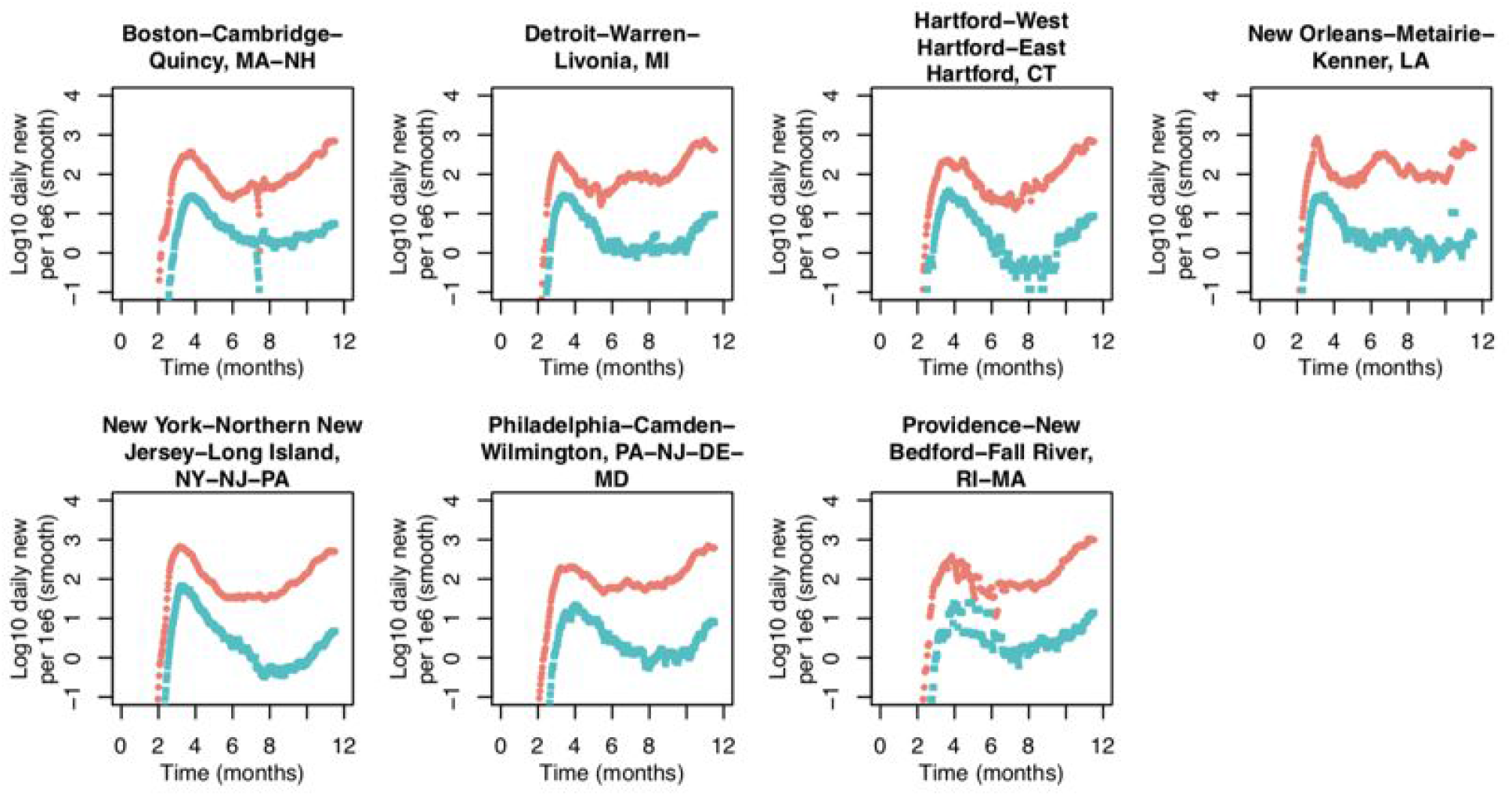
Cities following “Trajectory 1” of COVID-19 cases and deaths. Daily incidence of cases and deaths (per million, 7-day rolling average) in US metropolitan statistical areas that were assigned to the “Trajectory 1” group by hierarchical clustering of the time series. Note that the anomalously low cases in mid August in the Boston area are due to an approximately one week lapse in reporting at the county level in Massachusetts. The irregularities in the Providence area data are due to irregular county-level reporting in Rhode Island.

**Figure S6:**
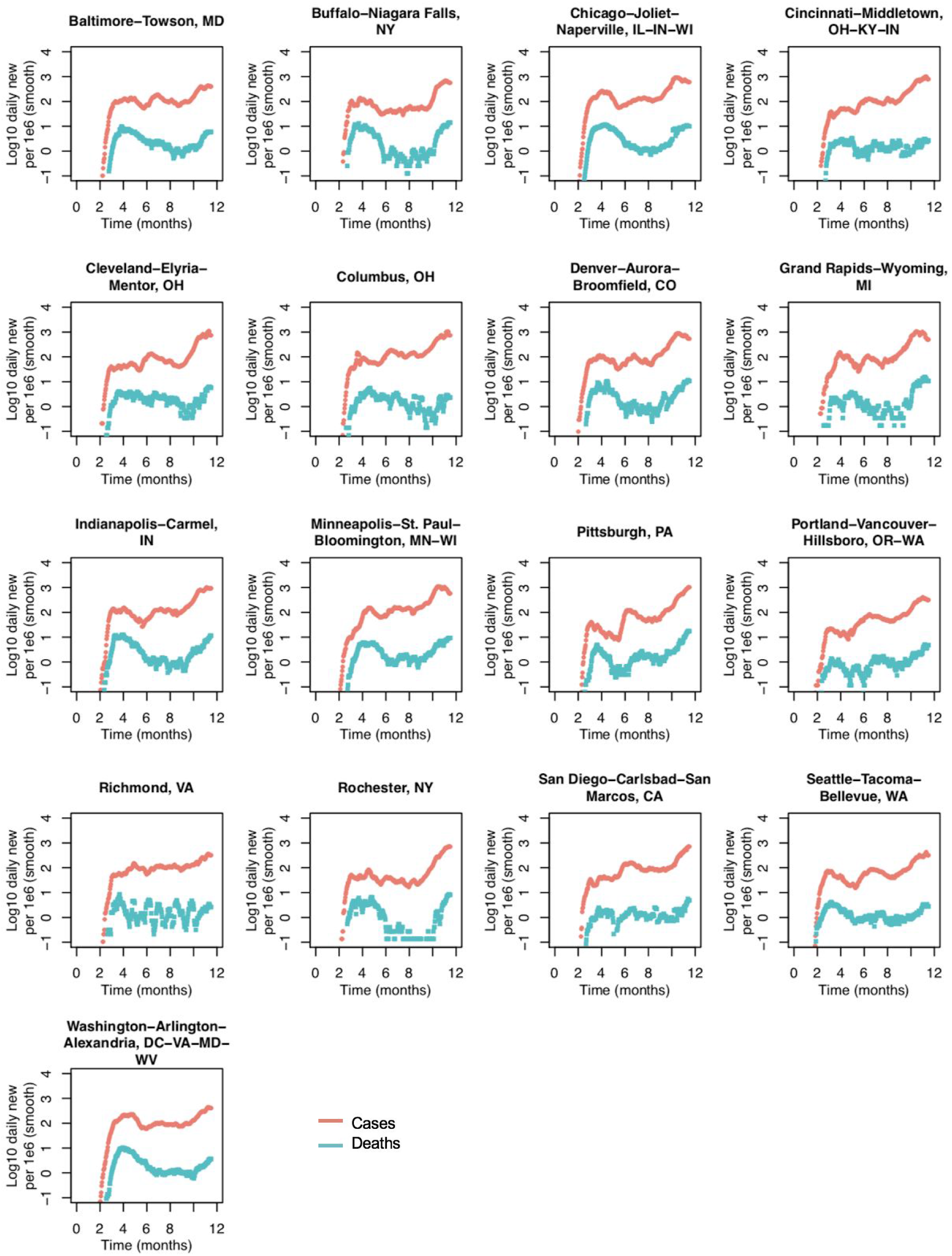
Cities following “Trajectory 2” of COVID-19 cases and deaths. Daily incidence of cases and deaths (per million, 7-day rolling average) in US metropolitan statistical areas that were assigned to the “Trajectory 2” group by hierarchical clustering of the time series.

**Figure S7:**
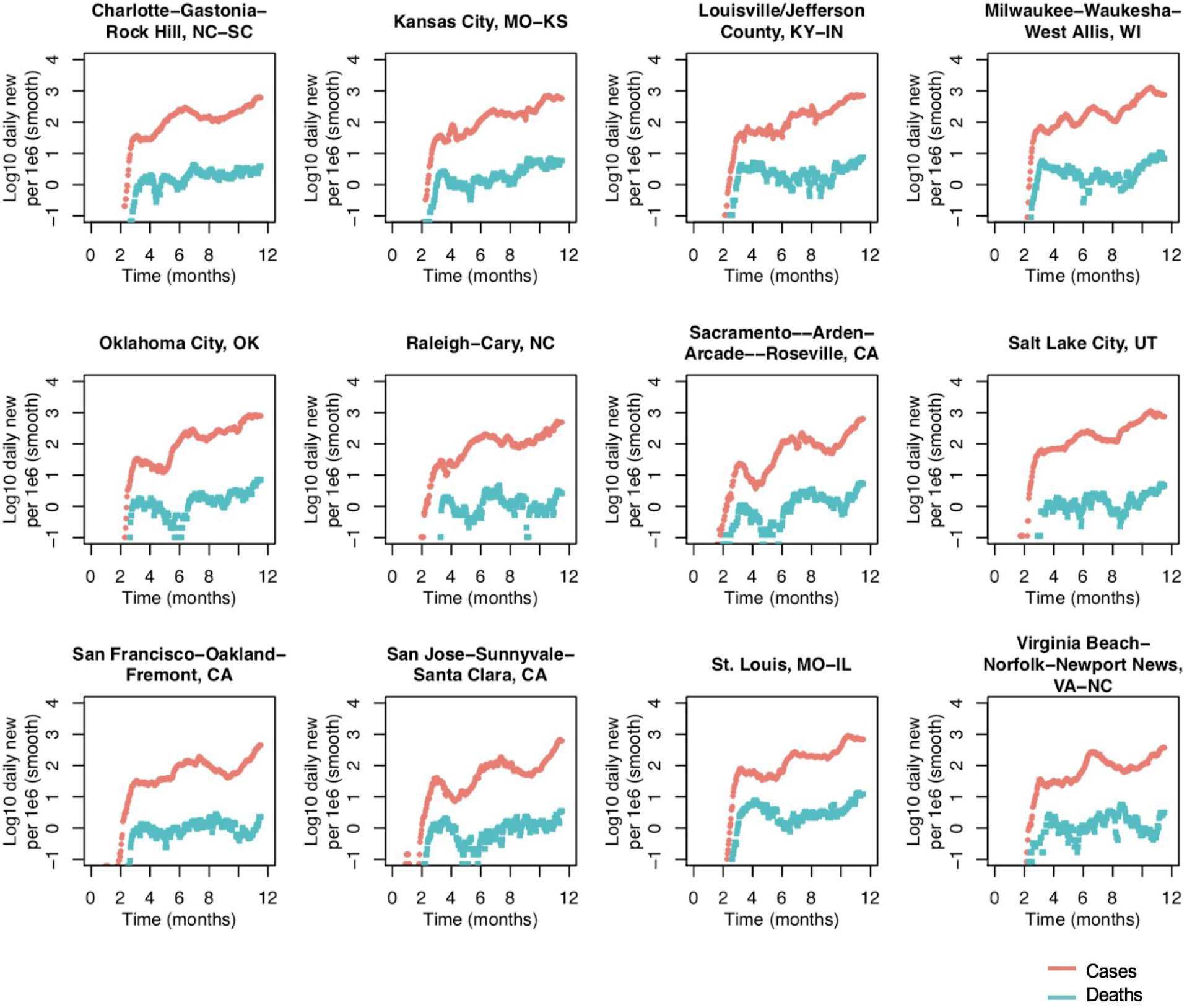
Cities following “Trajectory 3” of COVID-19 cases and deaths. Daily incidence of cases and deaths (per million, 7-day rolling average) in US metropolitan statistical areas that were assigned to the “Trajectory 3” group by hierarchical clustering of the time series.

**Figure S8:**
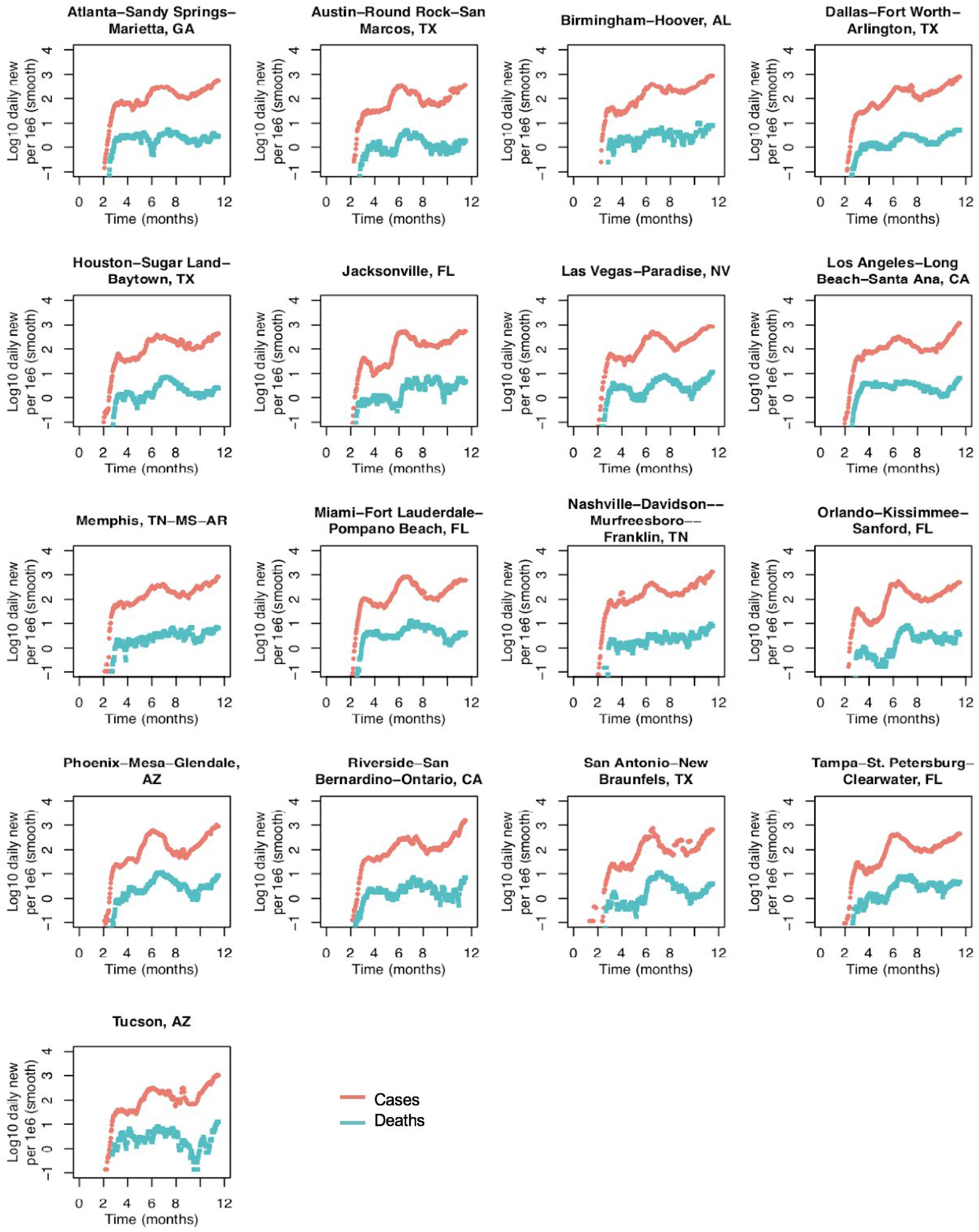
Cities following “Trajectory 4” of COVID-19 cases and deaths. Daily incidence of cases and deaths (per million, 7-day rolling average) in US metropolitan statistical areas that were assigned to the “Trajectory 4” group by hierarchical clustering of the time series.

**Figure S9:**
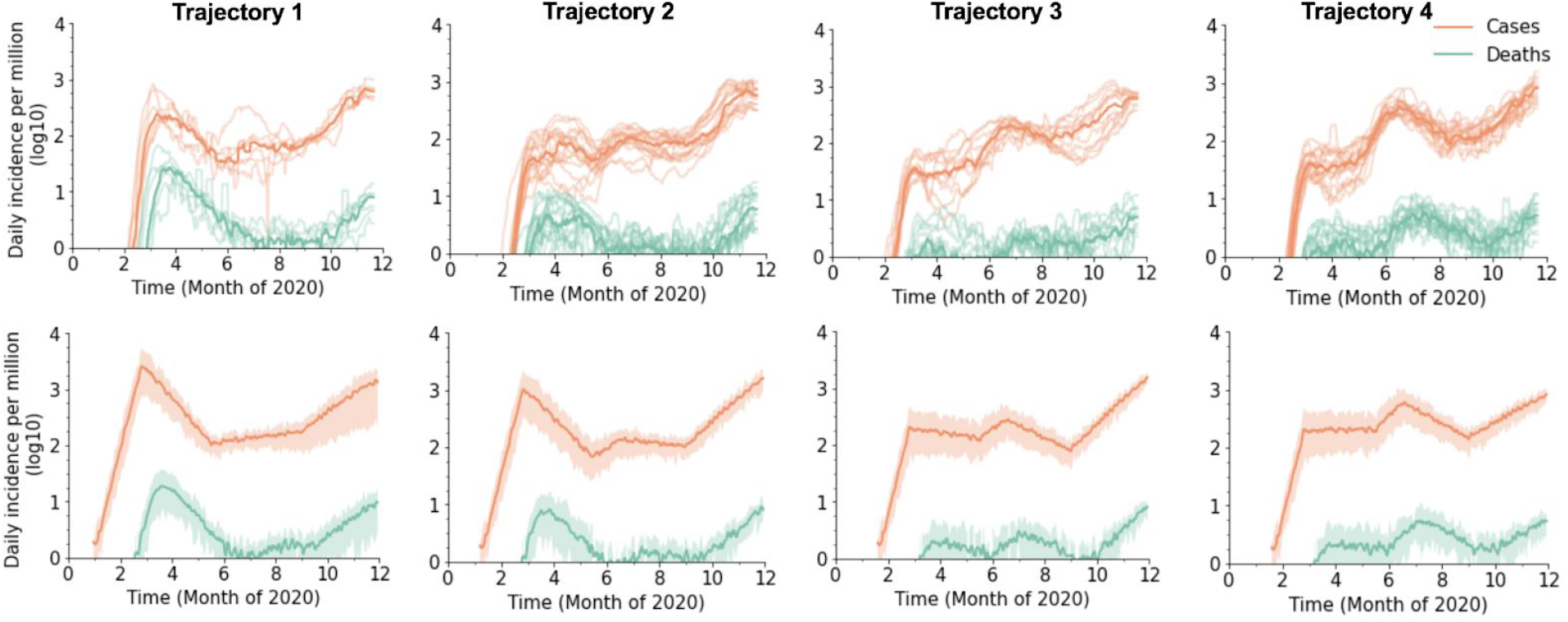
Calibrated model trajectories for each group of cities. All panels show daily incidence of new cases and deaths per million individuals, applying a 7-day rolling average. Top row: Case and death data for each metropolitan area assigned to each trajectory group by the clustering algorithm (see Figure S3). along with the group median (bolded line). Bottom row: Simulated case and death values from model scenarios calibrated to each group. Each simulated trajectory consists of up to five distinct phases of the epidemic wherein the number of external contacts is reduced by a fixed amount. The phases are the early exponential phase (pre March 25), the spring period of control/lockdown (March 25 - June 15), a relaxation of controls and possible summer resurgence (June 15 - July 15), an optional re-imposition of some controls (July 15 - Oct 1), and a fall comeback (Oct 1 onwards). Parameters describing each trajectory are given in Table S1. Note that in the data, cases are likely under-reported, especially during early phases of the epidemic, whereas the simulation counts all infections, so produces a higher ratio of cases to deaths.

**Figure S10:**
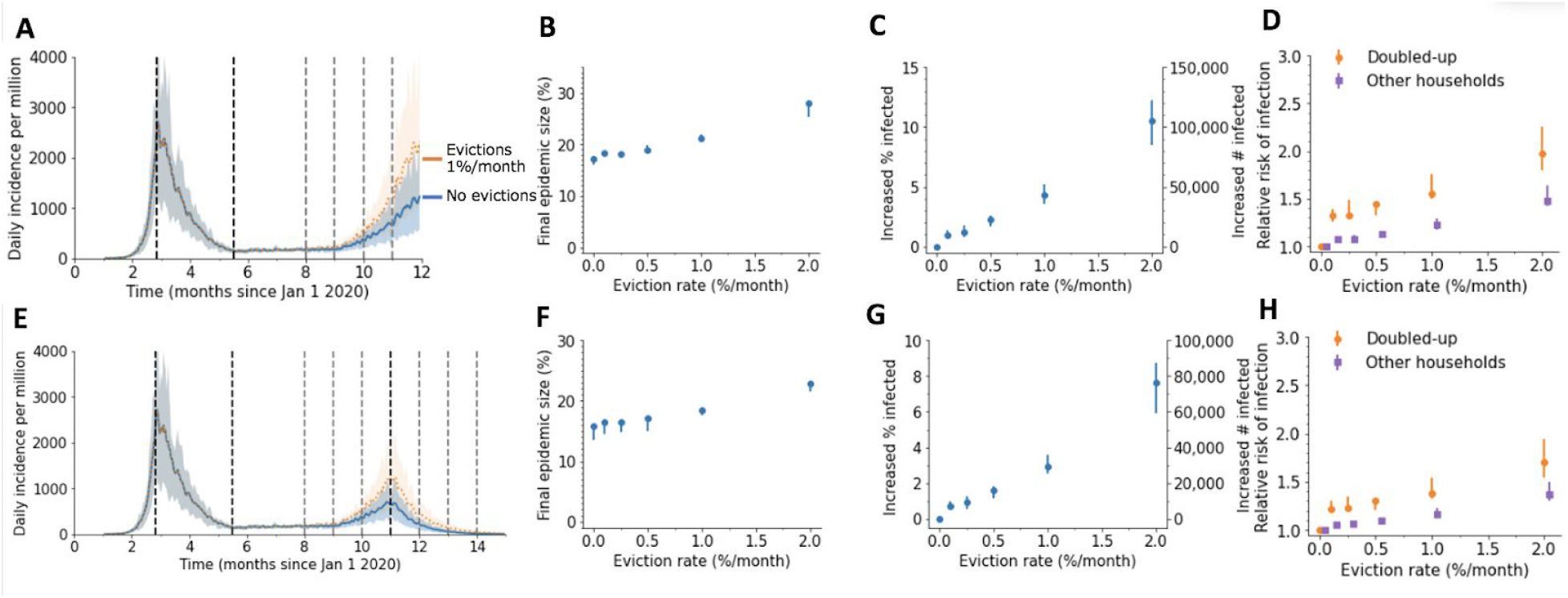
Impact of evictions on a SARS-CoV-2 comeback during Fall 2020 for a higher household SAR. We model evictions occurring in the context of an epidemic similar to cities following “Trajectory 1”, with a large first wave and control in the spring, followed by relaxation to a plateau over the summer and an eventual comeback in the fall. Monthly evictions start Sept 1, with a 4-month backlog processed on the first month. The household secondary attack rate (SAR) was 0.5. A) The projected daily incidence of new infections (7-day running average) with and without evictions. Shaded regions represent central 90% of all simulations.The first lockdown (dotted vertical line) reduced external contacts by 85%, under relaxation (second dotted line) they were still reduced by 70%, and during the fall comeback they were reduced by 60% (fourth dotted line). B) Final epidemic size by Dec 31 2020, measured as percent of individuals who had ever been in any stage of infection. C) The predicted increase in infections due to evictions through Dec 31 2020, measured as excess percent of population infected (left Y-axis) or number of excess infections (right Y-axis). Error bars show interquartile ranges across simulations. D) Relative risk of infection in the presence versus absence of evictions, for individuals who merged households due to evictions (“Doubled-up”) and for individuals who kept their pre-epidemic household (“Other households”). E)-F) Same as above but assuming a second lockdown is instituted on Dec 1, and maintained through March 2021.

**Figure S11:**
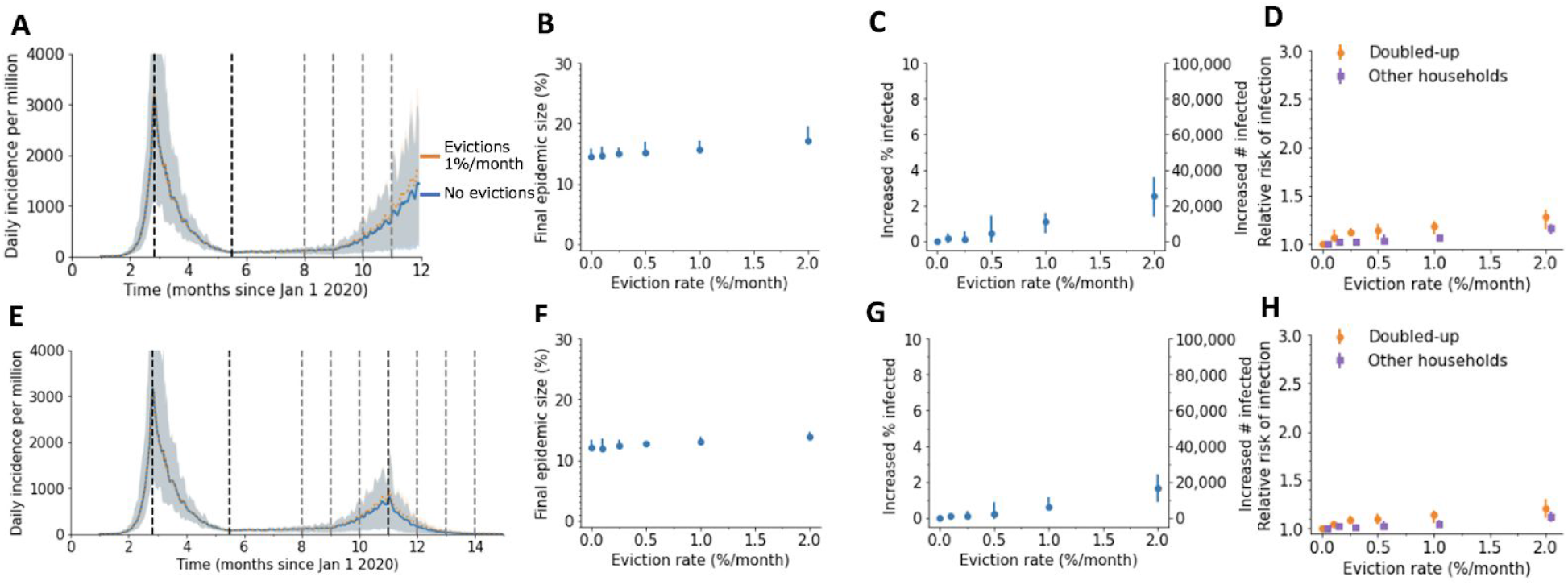
Impact of evictions on a SARS-CoV-2 comeback during Fall 2020 for a lower household SAR. We model evictions occurring in the context of an epidemic similar to cities following “Trajectory 1”, with a large first wave and control in the spring, followed by relaxation to a plateau over the summer and an eventual comeback in the fall. Monthly evictions start Sept 1, with a 4-month backlog processed on the first month. The household secondary attack rate (SAR) was 0.1. A) The projected daily incidence of new infections (7-day running average) with and without evictions. Shaded regions represent central 90% of all simulations.The first lockdown (dotted vertical line) reduced external contacts by 85%, under relaxation (second dotted line) they were still reduced by 70%, and during the fall comeback they were reduced by 60% (fourth dotted line). B) Final epidemic size by Dec 31 2020, measured as percent of individuals who had ever been in any stage of infection. C) The predicted increase in infections due to evictions through Dec 31 2020, measured as excess percent of population infected (left Y-axis) or number of excess infections (right Y-axis). Error bars show interquartile ranges across simulations. D) Relative risk of infection in the presence versus absence of evictions, for individuals who merged households due to evictions (“Doubled-up”) and for individuals who kept their pre-epidemic household (“Other households”). E)-F) Same as above but assuming a second lockdown is instituted on Dec 1, and maintained through March 2021.

**Figure S12:**
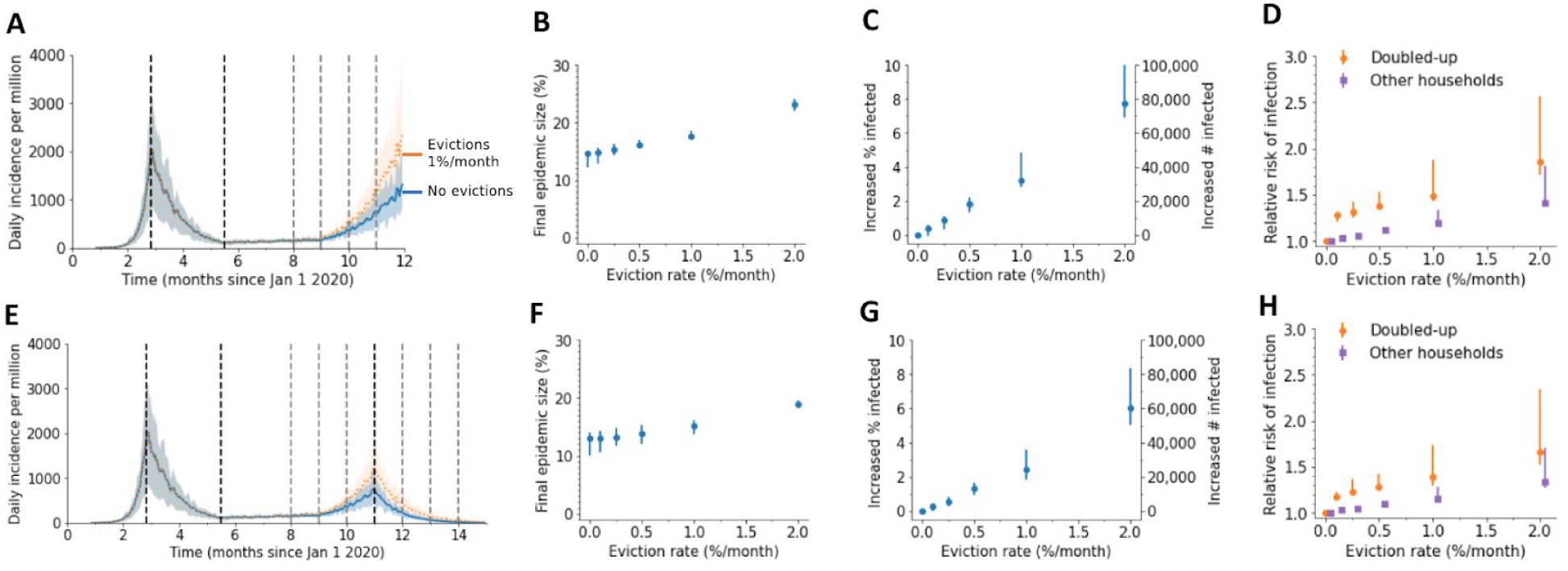
Impact of evictions on a SARS-CoV-2 comeback during Fall 2020 when household and external contacts have equal transmission risk. We model evictions occurring in the context of an epidemic similar to cities following “Trajectory 1”, with a large first wave and control in the spring, followed by relaxation to a plateau over the summer and an eventual comeback in the fall. Monthly evictions start Sept 1, with a 4-month backlog processed on the first month. A) The projected daily incidence of new infections (7-day running average) with and without evictions. Shaded regions represent central 90% of all simulations.The first lockdown (dotted vertical line) reduced external contacts by 85%, under relaxation (second dotted line) they were still reduced by 70%, and during the fall comeback they were reduced by 60% (fourth dotted line). B) Final epidemic size by Dec 31 2020, measured as percent of individuals who had ever been in any stage of infection. C) The predicted increase in infections due to evictions through Dec 31 2020, measured as excess percent of population infected (left Y-axis) or number of excess infections (right Y-axis). Error bars show interquartile ranges across simulations. D) Relative risk of infection in the presence versus absence of evictions, for individuals who merged households due to evictions (“Doubled-up”) and for individuals who kept their pre-epidemic household (“Other households”). E)-F) Same as above but assuming a second lockdown is instituted on Dec 1, and maintained through March 2021.

**Figure S13:**
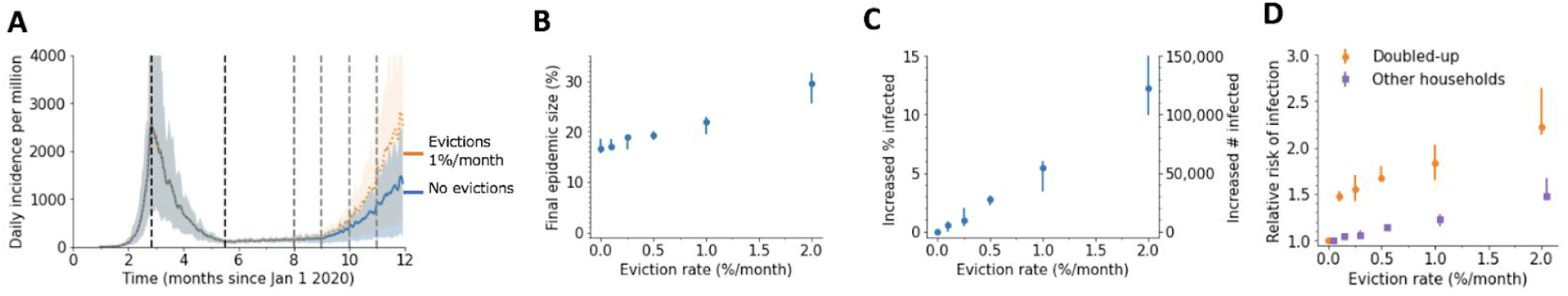
Impact of evictions on a SARS-CoV-2 comeback during Fall 2020 when doubling up occurs with a connected household. We model evictions occurring in the context of an epidemic similar to cities following “Trajectory 1”, with a large first wave and control in the spring, followed by relaxation to a plateau over the summer and an eventual comeback in the fall. Monthly evictions start Sept 1, with a 4-month backlog processed on the first month. A) The projected daily incidence of new infections (7-day running average) with and without evictions. Shaded regions represent central 90% of all simulations.The first lockdown (dotted vertical line) reduced external contacts by 85%, under relaxation (second dotted line) they were still reduced by 70%, and during the fall comeback they were reduced by 60% (fourth dotted line). B) Final epidemic size by Dec 31 2020, measured as percent of individuals who had ever been in any stage of infection. C) The predicted increase in infections due to evictions through Dec 31 2020, measured as excess percent of population infected (left Y-axis) or number of excess infections (right Y-axis). Error bars show interquartile ranges across simulations. D) Relative risk of infection in the presence versus absence of evictions, for individuals who merged households due to evictions (“Doubled-up”) and for individuals who kept their pre-epidemic household (“Other households”).

**Figure S14:**
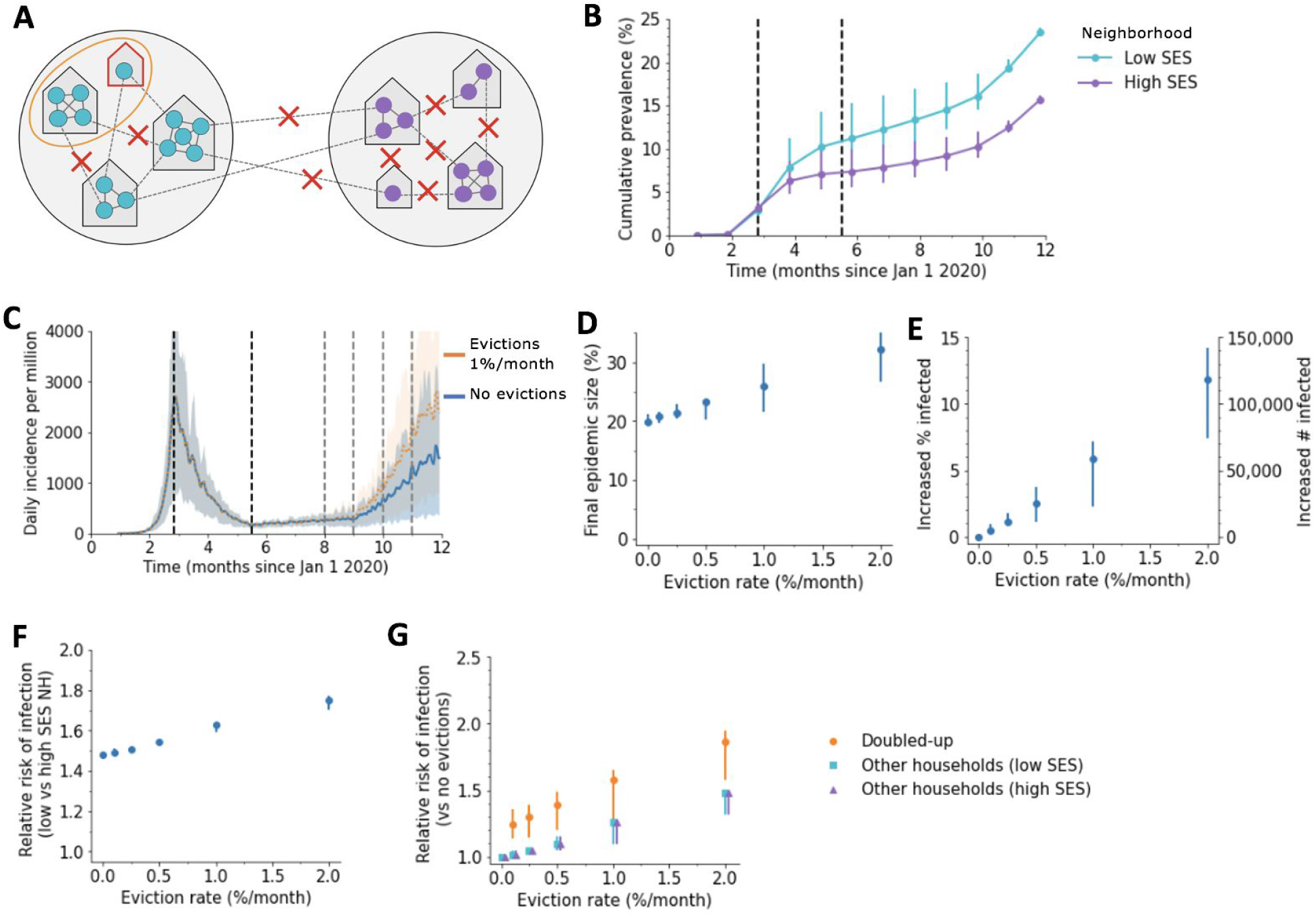
Impact of evictions on COVID-19 epidemics in heterogeneous cities with preferential mixing. A) Schematic of our model for inequalities within a city. The city is divided into a “high socioeconomic status (SES)” (purple) and a “low SES” (teal) neighborhood. Evictions only occur in the low SES area, and individuals living in this area are assumed to be less able to adopt social distancing measures, and hence have higher contact rates under interventions (90% vs 80% reduction in external contacts during lockdown for 85% overall, 75% vs 65% during relaxation for 70% overall, and 65% vs 55% during fall comeback for 60%). Before interventions, residents are more likely (75% external contacts are within one’s neighborhood) to contact someone outside the household who lives within vs outside their neighborhood. B) Cumulative percent of the population infected over time, by neighborhood, in the absence of evictions. C) The projected daily incidence of new infections (7-day running average) with 1%/month evictions vs no evictions. Shaded regions represent central 90% of all simulations. D) Final epidemic size by Dec 31 2020, measured as percent individuals who had ever been in any stage of infection, for the heterogenous city as compared to a homogenous city with same effective eviction rate and intervention efficacy. E) The predicted increase in infections due to evictions through Dec 31 2020, measured as excess percent of population infected (left Y-axis) or number of excess infections (right Y-axis). Error bars show interquartile ranges across simulations. F) Relative risk of infection by Dec 31 2020 for residents of the poor vs rich neighborhood. G) Relative risk of infection by Dec 31 2020 in the presence vs absence of evictions, for individuals who merged households due to evictions (“Doubled-up”) and for individuals who kept their pre-epidemic household (“Other households”).

**Figure S15:**
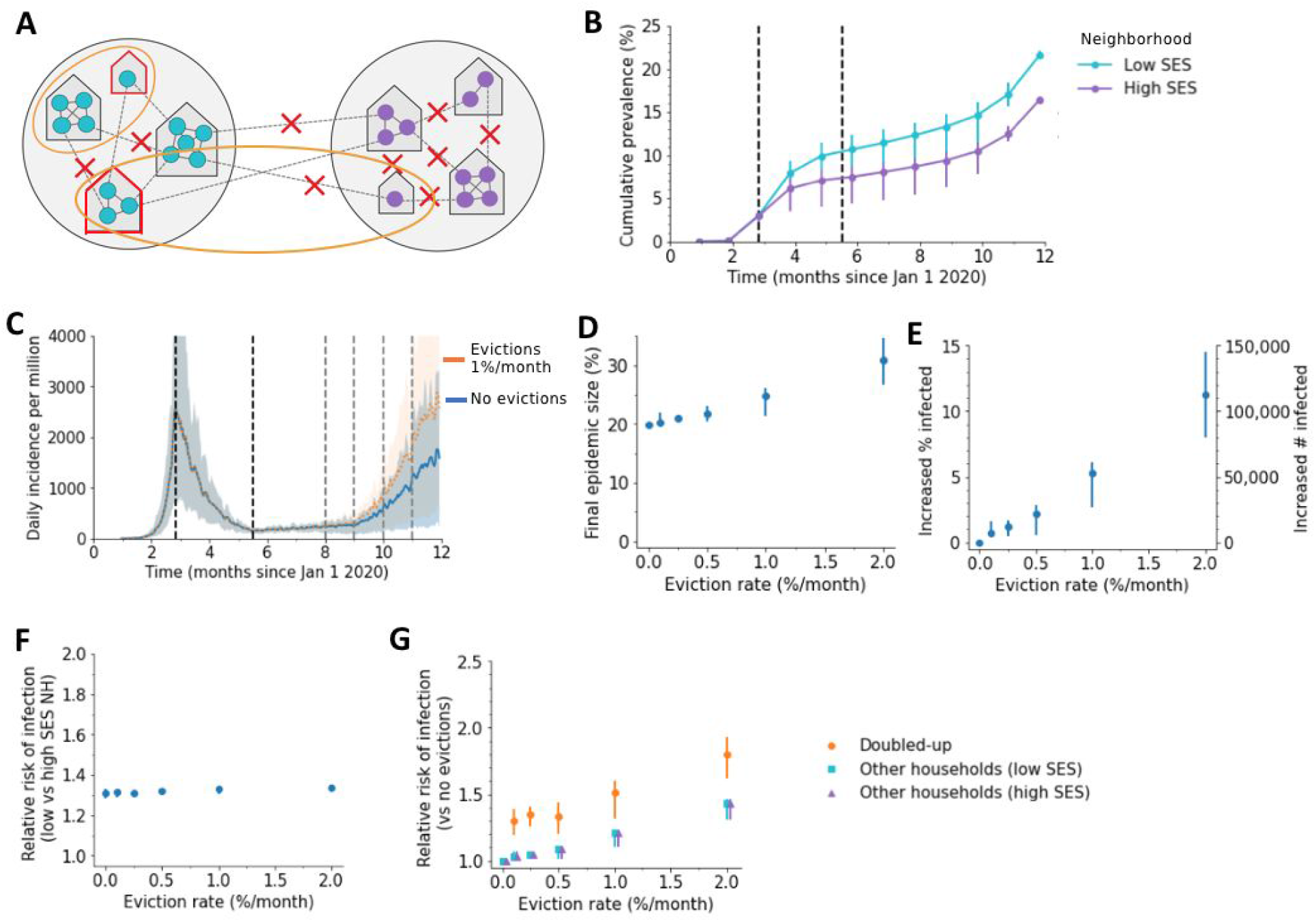
Impact of evictions on COVID-19 epidemics in heterogeneous cities with cross-neighborhood doubling up. A) Schematic of our model for inequalities within a city. The city is divided into a “high socioeconomic status (SES)” (purple) and a “low SES” (teal) neighborhood. Evictions only occur in the low SES area (though doubling up is equally likely to happen with individuals in any area), and individuals living in the low SES area are assumed to be less able to adopt social distancing measures, and hence have higher contact rates under interventions (90% vs 80% reduction in external contacts during lockdown for 85% overall, 75% vs 65% during relaxation for 70% overall, and 65% vs 55% during fall comeback for 60% overall). Before interventions, residents are equally likely to contact someone outside the household who lives within vs outside their neighborhood. B) Cumulative percent of the population infected over time, by neighborhood, in the absence of evictions. C) The projected daily incidence of new infections (7-day running average) with 1%/month evictions vs no evictions. Shaded regions represent central 90% of all simulations. D) Final epidemic size by Dec 31 2020, measured as percent individuals who had ever been in any stage of infection, for the heterogenous city as compared to a homogenous city with same effective eviction rate and intervention efficacy. E) The predicted increase in infections due to evictions through Dec 31 2020, measured as excess percent of population infected (left Y-axis) or number of excess infections (right Y-axis). Error bars show interquartile ranges across simulations. F) Relative risk of infection by Dec 31 2020 for residents of the poor vs rich neighborhood. G) Relative risk of infection by Dec 31 2020 in the presence vs absence of evictions, for individuals who merged households due to evictions (“Doubled-up”) and for individuals who kept their pre-epidemic household (“Other households”).

**Figure S16:**
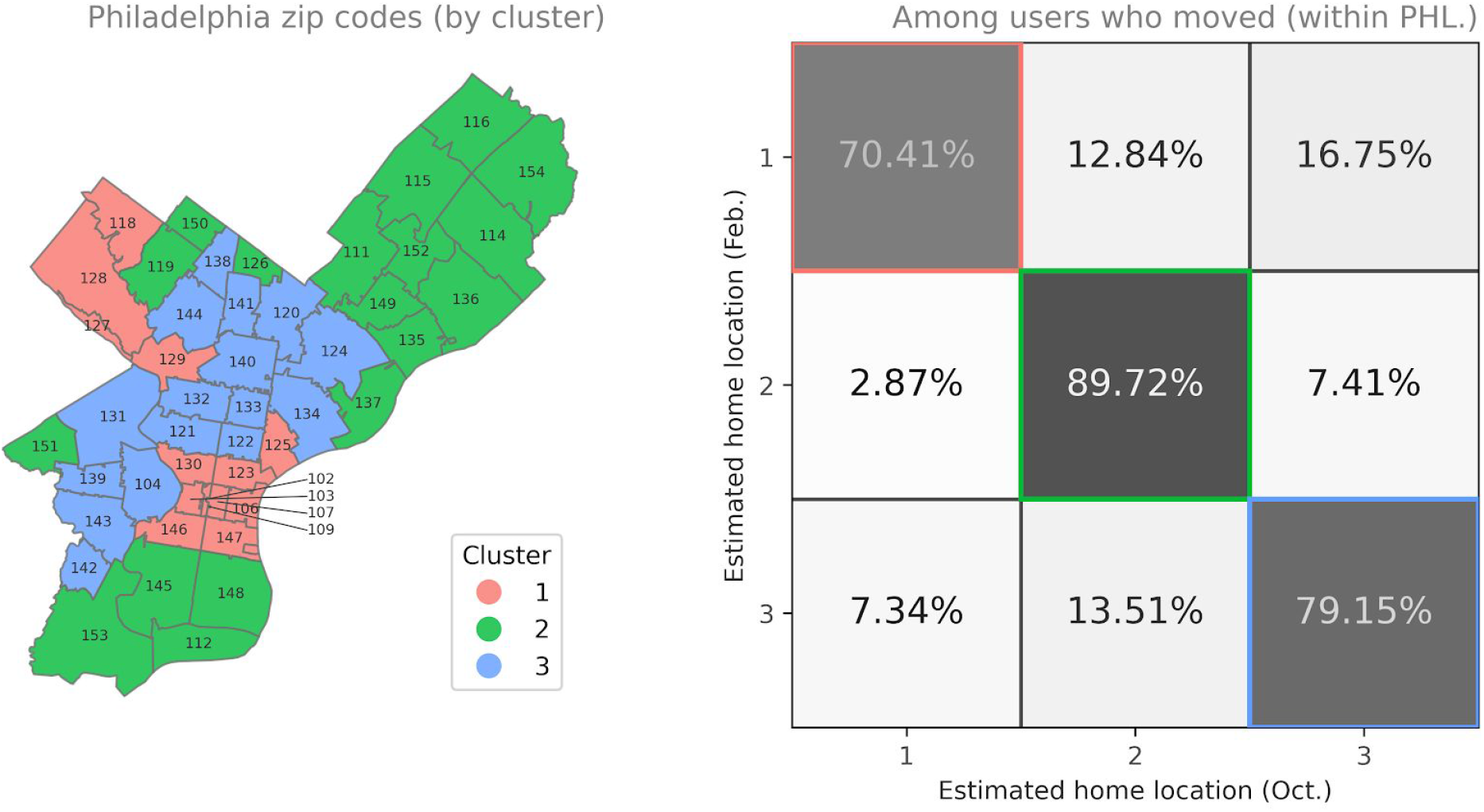
Relocation probabilities between clusters. Left: Map of Philadelphia, with each zip code colored by the cluster it was assigned to. Right: Among users in our panel who have a different estimated home location between February and October 2020, this matrix shows the percent of users who relocated from/to each cluster. Note that most moves took place within the same cluster.

**Figure S17:**
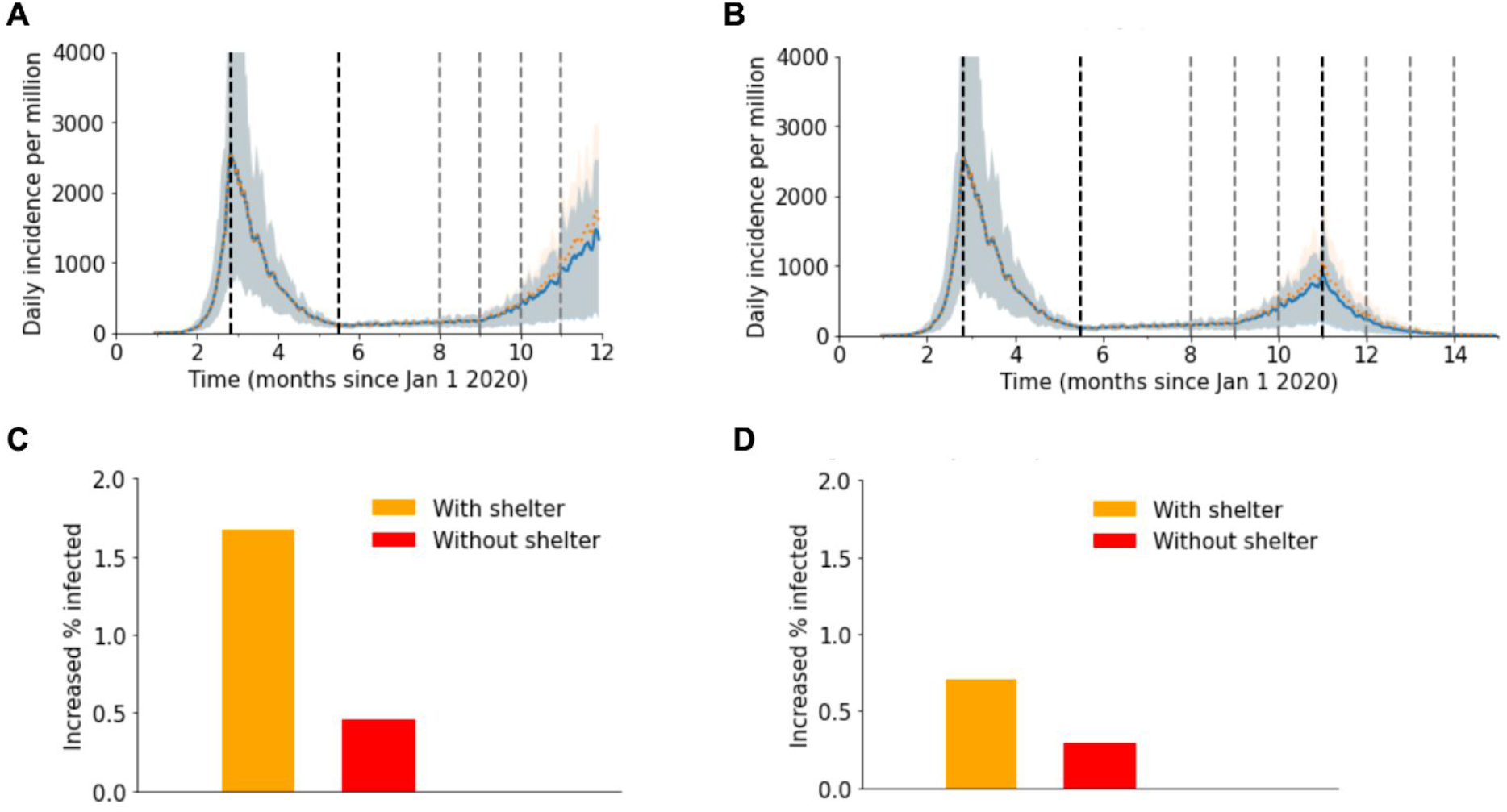
Impact of evictions on a SARS-CoV-2 when evictions can lead to homelessness. We model evictions occurring in the context of an epidemic similar to cities following “Trajectory 1”, with a large first wave and control in the spring, followed by relaxation to a plateau over the summer and an eventual comeback in the fall. Monthly evictions start Sept 1, with a 4-month backlog processed on the first month. Each month, 10% of evicted individuals become homeless and are assigned to a shelter, while the remaining 90% double-up. Each shelter size is drawn from a Poisson distribution with mean = 20 individuals. A)-B) The projected daily incidence of new infections (7-day running average) with (0.25% households per month) and without evictions. Shaded regions represent central 90% of all simulations.The first lockdown (dotted vertical line) reduced external contacts by 85%, under relaxation (second dotted line) they were reduced by 70%, and under the fall comeback they were reduced by 60% (fourth dotted line). In B) a second lockdown is imposed on Dec 1 and maintained through March 2021. C)-D) The predicted increase in infections through C) Dec 31, 2020 and D) March 31, 2021 due to evictions with (orange) and without (red) the inclusion of shelters for each of the above scenarios. It is measured in terms of an excess percent of the population infected.

## Supplemental Tables

**Table S1:** Parameters used for each simulated epidemic trajectory.

| | Base-line $R_0$ | Cumulative prevalence at lockdown (%) | Reduction in external contacts (%) | | | | |
| --- | --- | --- | --- | --- | --- | --- | --- |
|  |  |  | Early phase (pre March 25) | Lockdown (March 25) | Relaxation (June 15) | Control (July 15) | Comeback (Oct 1) |
| Trajectory Group (Figures 2, 3) |  |  |  |  |  |  |  |
| 1 | 3 | 3 | 0 | 85 | 70 | 70 | 60 |
| 2 | 3 | 1 | 0 | 85 | 65 | 75 | 60 |
| 3 | 3 | 0.2 | 0 | 77 | 65 | 80 | 60 |
| 4 | 3 | 0.2 | 0 | 75 | 60 | 80 | 65 |
| Sensitivity analyses (Figures S10-S12) |  |  |  |  |  |  |  |
| High SAR | 3.5 | 3 | 0 | 87 | 75 | 75 | 65 |
| Low SAR, Equal Weight | 3 | 3 | 0 | 80 | 65 | 65 | 55 |
| Equal Weight | 3 | 3 | 0 | 85 | 70 | 70 | 60 |
| Two-Neighborhood |  |  |  |  |  |  |  |
| Low-SES | 3 | 3 | 0 | 80 | 65 | 65 | 55 |
| High-SES | 3 | 3 | 0 | 90 | 75 | 75 | 65 |

**Table S2:**
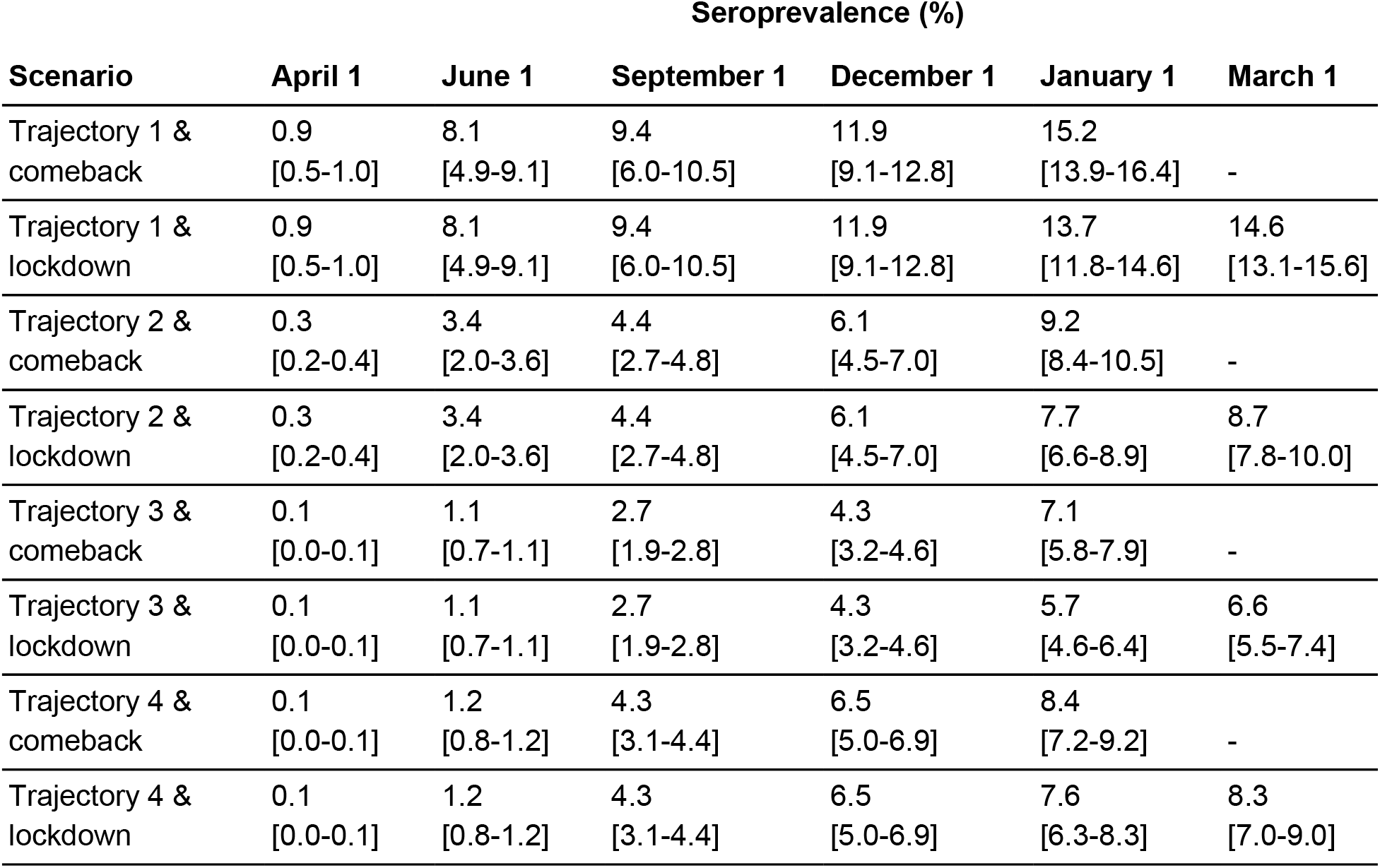
Seroprevalence over time in simulated epidemic trajectories.

| Scenario | Seroprevalence (%) |  |  |  |  |  |
| --- | --- | --- | --- | --- | --- | --- |
|  | April 1 | June 1 | September 1 | December 1 | January 1 | March 1 |
| Trajectory 1 & comeback | 0.9<br>[0.5-1.0] | 8.1<br>[4.9-9.1] | 9.4<br>[6.0-10.5] | 11.9<br>[9.1-12.8] | 15.2<br>[13.9-16.4] | - |
| Trajectory 1 & lockdown | 0.9<br>[0.5-1.0] | 8.1<br>[4.9-9.1] | 9.4<br>[6.0-10.5] | 11.9<br>[9.1-12.8] | 13.7<br>[11.8-14.6] | 14.6<br>[13.1-15.6] |
| Trajectory 2 & comeback | 0.3<br>[0.2-0.4] | 3.4<br>[2.0-3.6] | 4.4<br>[2.7-4.8] | 6.1<br>[4.5-7.0] | 9.2<br>[8.4-10.5] | - |
| Trajectory 2 & lockdown | 0.3<br>[0.2-0.4] | 3.4<br>[2.0-3.6] | 4.4<br>[2.7-4.8] | 6.1<br>[4.5-7.0] | 7.7<br>[6.6-8.9] | 8.7<br>[7.8-10.0] |
| Trajectory 3 & comeback | 0.1<br>[0.0-0.1] | 1.1<br>[0.7-1.1] | 2.7<br>[1.9-2.8] | 4.3<br>[3.2-4.6] | 7.1<br>[5.8-7.9] | - |
| Trajectory 3 & lockdown | 0.1<br>[0.0-0.1] | 1.1<br>[0.7-1.1] | 2.7<br>[1.9-2.8] | 4.3<br>[3.2-4.6] | 5.7<br>[4.6-6.4] | 6.6<br>[5.5-7.4] |
| Trajectory 4 & comeback | 0.1<br>[0.0-0.1] | 1.2<br>[0.8-1.2] | 4.3<br>[3.1-4.4] | 6.5<br>[5.0-6.9] | 8.4<br>[7.2-9.2] | - |
| Trajectory 4 & lockdown | 0.1<br>[0.0-0.1] | 1.2<br>[0.8-1.2] | 4.3<br>[3.1-4.4] | 6.5<br>[5.0-6.9] | 7.6<br>[6.3-8.3] | 8.3<br>[7.0-9.0] |

**Table S3:** Socioeconomic indicators used to classify zipcodes in Philadelphia into clusters.

| Indicator Description | Mean by Cluster |  |  |  |
| --- | --- | --- | --- | --- |
|  | Overall mean | Cluster 1 | Cluster 2 | Cluster 3 |
| Population under 18 (%) | 19.8% | 11.4% | 21.6% | 24.5% |
| Population over 65 (%) | 13.8% | 13.7% | 15.9% | 11.4% |
| Female (%) | 52.4% | 51.1% | 52.4% | 53.5% |
| Poverty Rate (%) | 22.9% | 14.4% | 17.6% | 36.1% |
| Per Capital Income (\$) | 31,436 | 54,996 | 27,250 | 17,265 |
| Median Household Income (\$) | 50,391 | 75,828 | 50,414 | 29,695 |
| Median Home Value (\$) | 198,396 | 353,577 | 177,100 | 97,600 |
| Renter Occupied (%) | 47.6% | 55.7% | 37.5% | 52.9% |
| Vacancy Rate (%) | 12.6% | 11.9% | 9.2% | 17.0% |
| Median Gross Rent (\$) | 1,092 | 1,448 | 1,023 | 885 |
| Housing Cost Burdened Owners (%) | 27.9% | 25.1% | 27.4% | 30.8% |
| Housing Cost Burdened Renters (%) | 48.9% | 38.3% | 50.8% | 55.4% |
| Female Headed Households (%) | 18.8% | 7.7% | 18.5% | 28.1% |
| Residents Employed in Service Occupations (%) | 22.2% | 11.8% | 23.2% | 29.4% |
| Residents Employed in Essential Services (%) | 46.2% | 30.4% | 50.4% | 54.1% |
| Mobility Rate (% moved within last year) | 15.9% | 24.7% | 10.5% | 15.1% |
| Commute over 60 minutes (%) | 14.4% | 9.6% | 15.1% | 17.6% |
| Population (Thousands) | 1,584 | 269 | 602 | 713 |
| Proportion of Population (%) | 100% | 17% | 38% | 45% |

**Table S4:** Composition of zipcode-clusters in Philadelphia by race/ethnicity/nativity.

|  | **Cluster 1** | **Cluster 2** | **Cluster 3** |
| --- | --- | --- | --- |
| White (non-Hispanic) | 69.8% | 51.1% | 19.7% |
| Black | 16.1% | 34.1% | 65.2% |
| Pacific Islander/American Indian/Alaska Native | 0.2% | 0.3% | 0.4% |
| Asian | 8.8% | 7.8% | 4.2% |
| Non-white | 30.2% | 48.9% | 80.3% |
| Latino/Hispanic | 6.6% | 10.2% | 17.7% |
| Foreign Born | 11.5% | 16.7% | 10.1% |

**Table S5:**
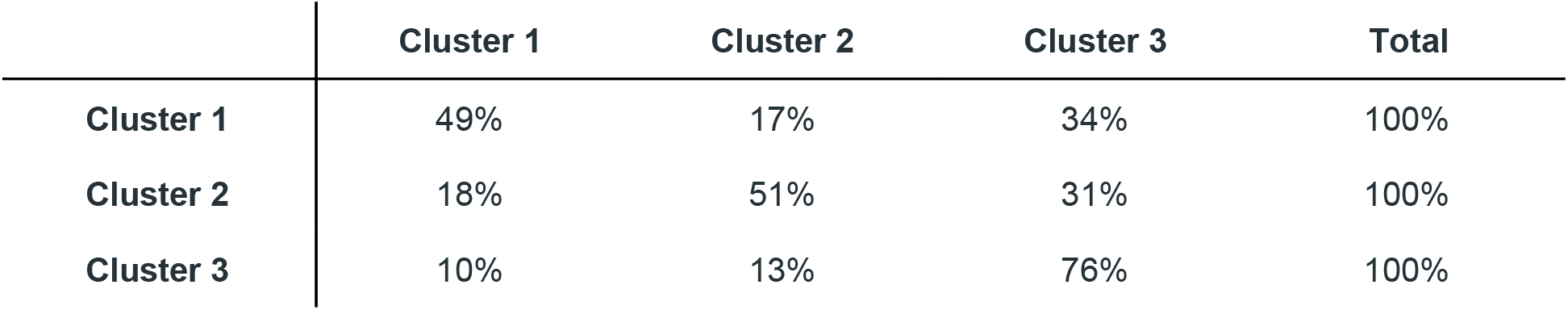
Baseline contact matrix by cluster of residence. Each row gives the fraction of all within-Philadelphia contacts involving individuals of a particular cluster that are with individuals residing in each other cluster. Contacts are estimated from co-location data from mobile devices (see Methods/Supplementary Methods). These mixing patterns were calculated for January and February 2020 and are used as the baseline patterns to which all COVID-19-driven reductions are applied.

**Table S6:** Reduction in contacts by clusters of residence and month. Each row gives the percent reduction in contacts between individuals belonging to different clusters as compared to the baseline contacts. Reductions are for the months of April (top line in each row) and June (bottom line in each row) and are estimated from co-location data from mobile devices (see Methods/Supplementary Methods).

|  | **Cluster 1** | **Cluster 2** | **Cluster 3** | **Net** |
| --- | --- | --- | --- | --- |
| **Cluster 1** | 94% | 99% | 99% | 96% |
|  | 85% | 91% | 94% | 89% |
| **Cluster 2** | 98% | 83% | 90% | 88% |
|  | 88% | 64% | 77% | 73% |
| **Cluster 3** | 98% | 92% | 91% | 92% |
|  | 92% | 79% | 82% | 83% |

